# Primary care coding activity related to the use of online consultation systems or remote consulting: an analysis of 53 million peoples’ health records using OpenSAFELY

**DOI:** 10.1101/2023.01.25.23284428

**Authors:** Martina Fonseca, Brian MacKenna, Amir Mehrkar, The OpenSAFELY Collaborative, Caroline E Walters, George Hickman, Jonathan Pearson, Louis Fisher, Peter Inglesby, Seb Bacon, Simon Davy, William Hulme, Ben Goldacre, Ofra Koffman, Minal Bakhai

## Abstract

**Background:** The pandemic accelerated work by the NHS in England to enable and stimulate use of online consultation systems across all practices, for improved access to primary care.

**Objective:** We aimed to explore general practice coding activity associated with the use of online consultation systems in terms of trends, COVID-19 effect, variation and quality.

**Methods:** With the approval of NHS England, OpenSAFELY-TPP and OpenSAFELY-EMIS were used to query and analyse *in situ* records of electronic health record systems of over 53 million patients in over 6,400 practices, mainly in 2019-2020. SNOMED CT codes relevant to online consultation systems and written online consultations were identified. Coded events were described by volumes, practice coverage, trends pre- and post-COVID-19 and inter-practice and sociodemographic variation.

**Results:** 3,550,762 relevant coding events were found in TPP practices, with code eConsultation detected in 84% of practices. Coding activity related to digital forms of interaction increased rapidly from March 2020 at the onset of the COVID-19 pandemic, though we found large variation in coding instance rates among practices in England. Code instances were more commonly found among females, those aged 18-40, those least deprived or white. eConsultation coded activity was more commonly found recorded among patients with a history of asthma or depression.

**Conclusions:** We successfully queried general practice coding activity relevant to the use of online consultation systems, showing increased adoption as well as key areas of variation during the COVID-19 pandemic. The work can be expanded to support monitoring of coding quality and underlying activity. In future, large-scale impact evaluation studies can be implemented within the platform, namely looking at resource utilisation and patient outcomes.

## Background

NHS England’s Digital First Primary Care (DFPC) programme^1^ has led the work on enabling the implementation and improvement of the use of digital tools in general practice, including the use of online consultation systems (OCS) and video consultations. The NHS Long Term plan^2^, published in January 2019 before the COVID-19 pandemic, committed to every patient having the right to digital-first primary care by 2023/24; the 5-year GP contract reform framework to support the NHS Long Term Plan, specifically committed for all practices to offer online and video consultation systems by 2021. Legislation that has come into force in October 2021 requires all NHS general practitioner (GP) practices to make an online consultation system available to their patients^3^. The COVID-19 pandemic response significantly accelerated the adoption and need for such systems. An internal NHS England data collection of information provided by OCS suppliers ^4^ rapidly stood up in April 2020 showed that as of 21st of June 2021 approximately 79% of the practices in England had online consultation system capability in place. This was believed to stand closer to 95% when considering gaps in data collection ^51^. In early 2021 there were about 10 weekly online consultation submissions per 1,000 population, though underlined by local variation.^2^

Given the rapid nationwide adoption and utilisation of online consultation systems and other digital tools (including alongside and in response to COVID-19), there is a need to better understand how they are: used, implemented, and generate impact. Monitoring and evaluation have been stood up by the Digital First Primary Care (DFPC) team, as well as independent studies. Namely, commissioned work includes research questions comparing different models of implementation and different types of OCS, analysis of patient experience and analysis of impact of digital tools on outcomes such as prescribing patterns, A&E attendances and emergency admissions ^6,7^. However, activity monitoring datasets such as the General Practice Appointment Data (GPAD) ^8^ or the internal NHS England data collection from OCS suppliers are aggregate and do not capture sociodemographics, clinical history or patient pathways. GPAD is classified as experimental data due to variations in practice coverage. The publication presents details of patient appointments that are recorded in the GP appointments system, rather than the totality of interactions. It cannot be considered to be a complete view of general practice activity. It does not currently capture all triage activity, all appointments following from online requests to practices from patients as they are often managed within other IT products nor does it currently include all enhanced access evening and weekend appointments which are managed by other appointment books. In terms of consultation appointment mode it only includes a broad ‘Video/online’ category, which is returned with variable completion and standardisation. On the other hand, more targeted pilots and evaluation studies have been designed that do access richer quantitative or qualitative information ^6,7,9,10^, but these tend to be localised to a single supplier or set of practices, so may not be nationally representative or provide a full picture along the pathway.

OpenSAFELY is a new secure analytics platform for electronic health records (EHR) in the NHS, created to deliver urgent insights during the global COVID-19 emergency. The platform uses a novel approach for enhanced security and timely data access that avoids the need to migrate large volumes of disclosive pseudonymised patient data outside of the secure environments managed by the EHR software companies (e.g. TPP, EMIS); instead, it relies on trusted analysts to run computations and analysis on near real-time pseudonymised patient records still held inside the data centres and secure cloud environments of EHR companies. With the approval of NHS England, we conducted a service evaluation using the NHS England OpenSAFELY COVID-19 research platform. In this particular study we explore EHR coding activity that is related to the use of online consultation systems (OCS) and online written consultations (that is, responses delivered by SMS or online messages) in general practices, via OpenSAFELY. By using primary care EHR system data, it is possible to leverage nationally representative, longitudinal patient cohort data regarding clinical and administrative encounters and to analyse this in the context of other factors such as geography, sociodemographics, clinical characteristics and other healthcare interactions. Given the limited pre-existing standardisation and insight into coding of submissions received via an online consultation system and coding of the mode of the consultation, we set out to focus on the following aims:

a. understanding online consultation system (OCS) and online written consultation **coding use and prevalence in primary care records** by codes of interest (as shorthand, ‘OCS-relevant’)
b. understanding the variation in OCS-relevant coding use over time, pre- and post-start of COVID-19 pandemic and in terms of inter-practice variation
c. understanding **broad demographics and past clinical history** of those with OCS-relevant coding activity.

This exploratory analysis may help inform further research and evaluation questions and their feasibility, including large-scale impact evaluation studies.

### Box 1

Online consultation systems; electronic health record systems

An **online consultation system** is an online facility that allows a patient or carer to seek advice or information related to the patient’s health, or to make a clinical or administrative request through completing an electronic form. The written information provided by patients or carers about the issue they are seeking help for enables practices to prioritise patient care based on clinical need and to ensure that care is offered by the right member of staff or service and in the right way. The mode of response is based on the clinical need, circumstances, and patient’s communication preferences.

While online consultations systems offer the patient or carer a way to access care, clinical staff decide (taking into account patients’ preferences) whether the patient will be responded to via a telephone call, invited to a video consultation, invited for a face-to-face appointment, or whether they will receive a **written response** in an electronic form e.g. SMS or online message ^34^.

Online consultation systems enable digital forms of patient-practice interaction and have only recently been more widely implemented.

The study also mentions **electronic health record (EHR) systems**. These are well established and are used as a much broader means to record and identify activity electronically, mainly through coding terminology. These contain results of clinical and administrative encounters between a provider (physician, nurse,, and others) and a patient that occur during episodes of patient care (it is not specific to recording digital forms of patient-practice interaction, though that would be in scope) ^35^.

## Methods

### Study design

We conducted a retrospective cohort study using general practice primary care EHR data from all GP practices in England with EHR vendors TPP and EMIS as suppliers. The present analysis project is part of a ‘Ways of working’ pilot to onboard into OpenSAFELY any new approved users or researchers (including NHS England analysts) ^11^.

### Data Source

All data were linked, stored and analysed securely within the OpenSAFELY platform (https://opensafely.org/), a data analytics platform created with the approval of NHS England to address urgent COVID-19 research questions. Data records used in this study are general practice primary care EHR data from practices in England that are supplied by the vendors TPP and EMIS. These contain data such as diagnoses, medications and sociodemographic characteristics.

Similarly pseudonymised datasets from other data providers are securely provided to the EHR vendor and linked to the primary care data, such information on care home status. No free text data are included. The TPP database analysed via OpenSAFELY (OpenSAFELY-TPP) is based on 24.2 million people currently registered with 2546 GP surgeries using TPP SystmOne software while the EMIS database analysed within OpenSAFELY (OpenSAFELY-EMIS) is based on 32.6 million people currently registered with 3821 GP surgeries using EMIS. Together, these represent about 99% of practices. Most of the outcomes in this study explore OpenSAFELY-TPP, though a more fixed-scope overall coding use and prevalence characterisation was extended to OpenSAFELY-EMIS as well, based on what was feasible in its early operational days (Supplementary Information).

OpenSAFELY-TPP and OpenSAFELY EMIS were used side-by-side to identify the use of online consultation relevant activity in the period surrounding the pandemic (January 2019 until December 2020) in terms of individual code utilisation and practice coverage. OpenSAFELY-TPP, covering about 40% of practices, was used to understand trends in coding activity over time (pre- and post-start of COVID-19 pandemic), inter-practice variation, associations with sociodemographic factors and associations with clinical history factors.

The queried pseudonymised records are available through the OpenSAFELY framework. Detailed pseudonymised patient data is potentially re-identifiable and therefore not shared. The codebase and aggregate non-disclosive outputs are available for use. Further details on information governance can be found under Information Governance and Ethics.

For benchmarking and triangulation, data from the national OC/VC supplier collection is also used. This data collection was stood up rapidly at the start of the pandemic and includes aggregate utilisation data taken directly from the participating OCS suppliers. ^1^ This contains daily information from August 2020, derived from the daily collection files. It can be extended as a weekly trend back to April 2020. No information on demographics, clinical history or pathway is part of its specifications. On completeness, an audit undertaken on the 31st of March 2021 suggested that approximately 10% of practices were using an online consultation supplier system that did not contribute to the national collection. As of September 2021, there were 5 out of 20 suppliers that were not submitting their data. This, however, has been evolving as several suppliers are working towards submitting their data.

### Coding systems

In general practice, staff record information about patients using clinical coding systems such as SNOMED CT and dm+d. System TPP is fully compliant with SNOMED, with GPs using it in their front-end interactions with EHR systems having previously used CTV3 before the NHS wide standard was adopted. OpenSAFELY can query the records using either CTV3 or SNOMED which allows flexibility on querying some past activity that cannot be easily mapped to SNOMED CT.

### Approach to deciding codes for interrogation

We could not ascertain the existence of a nationally consistent and standardised codelist for activity associated with online consultation systems, or with the carrying out of a written online consultation^3^, though work is underway to address this. This relates to a number of reasons including the recency of technology implementation and adoption, the lack of standardisation of terminology and appropriate codes around digital forms of interaction with a practice (by route and mode) and the disparity between supplier systems and templates. As such, a SNOMED CT codelist was created on OpenCodelists with existing relevant codes ^12^. This is available for inspection and re-use by anyone ^12^. The codes are also given in Table 1.

**Table 1.**
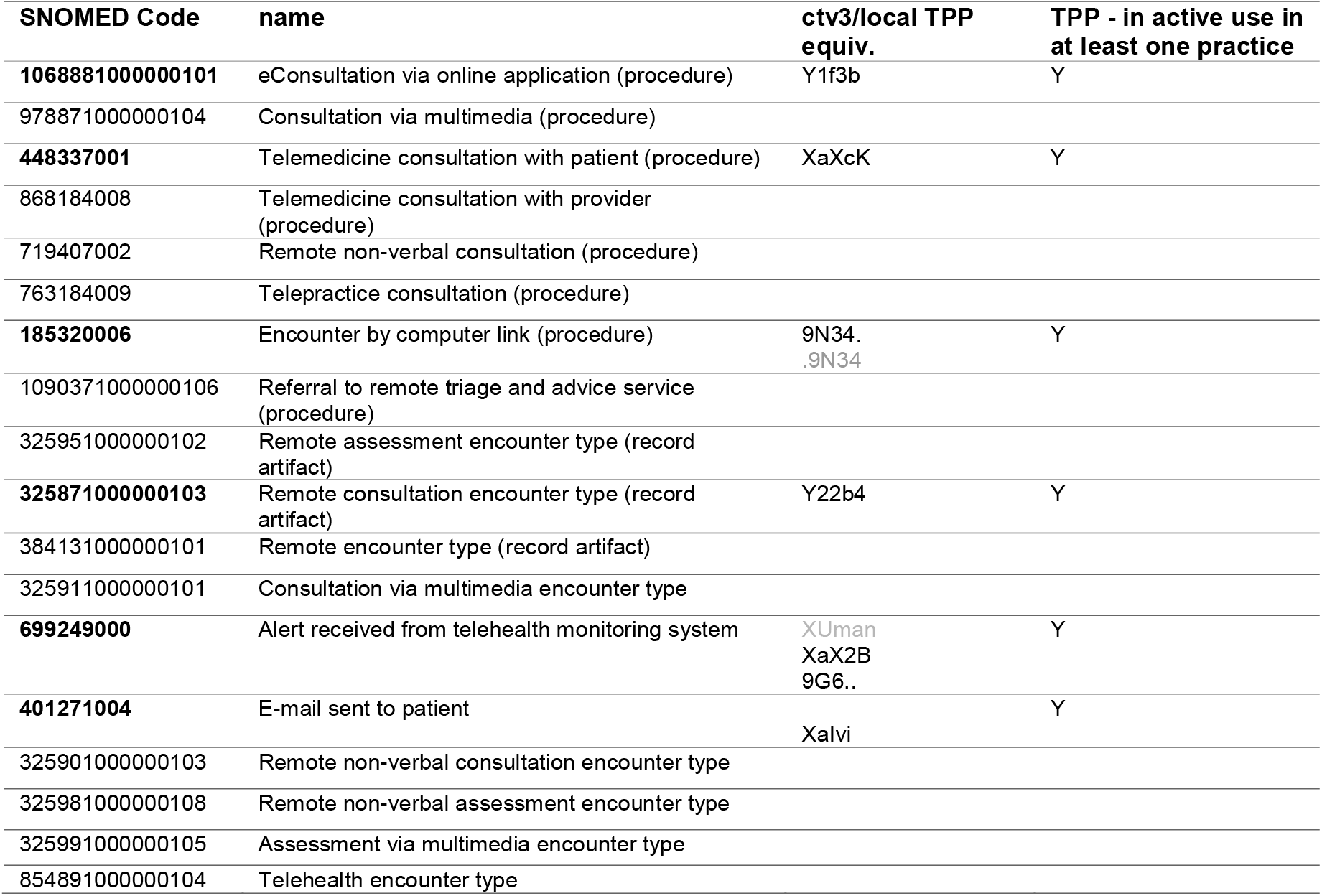
SNOMED shortlist: The short-listed read codes in SNOMED (codelist builder list available) ^12^

| SNOMED Code | name | ctv3/local TPP equiv. | TPP - in active use in at least one practice |
| --- | --- | --- | --- |
| **1068881000000101** | eConsultation via online application (procedure) | Y1f3b | Y |
| 978871000000104 | Consultation via multimedia (procedure) |  |  |
| **448337001** | Telemedicine consultation with patient (procedure) | XaXcK | Y |
| 868184008 | Telemedicine consultation with provider (procedure) |  |  |
| 719407002 | Remote non-verbal consultation (procedure) |  |  |
| 763184009 | Telepractice consultation (procedure) |  |  |
| **185320006** | Encounter by computer link (procedure) | 9N34.<br>.9N34 | Y |
| 1090371000000106 | Referral to remote triage and advice service (procedure) |  |  |
| 325951000000102 | Remote assessment encounter type (record artifact) |  |  |
| **325871000000103** | Remote consultation encounter type (record artifact) | Y22b4 | Y |
| 384131000000101 | Remote encounter type (record artifact) |  |  |
| 325911000000101 | Consultation via multimedia encounter type |  |  |
| **699249000** | Alert received from telehealth monitoring system | XUman<br>XaX2B<br>9G6.. | Y |
| **401271004** | E-mail sent to patient | Xalvi | Y |
| 325901000000103 | Remote non-verbal consultation encounter type |  |  |
| 325981000000108 | Remote non-verbal assessment encounter type |  |  |
| 325991000000105 | Assessment via multimedia encounter type |  |  |
| 854891000000104 | Telehealth encounter type |  |  |

In terms of criteria, the term ‘online consultations’ is ambiguous and can include submissions/requests received from patients via an online consultation system (route of access) or the mode of a consultation using written electronic messaging (appointment mode). Codes of interest were those deemed to be associated with either submissions made using an online consultation system (route) or written online consultations (mode). Keywords included in the search were: consultation (procedure); econsultation; indirect encounter; online; remote triage; telemedicine; telepractice

The codelist was developed as follows: a) browse the SNOMED CT Term Browser ^7^ for relevant keywords and children. Find its CTV3 equivalent (refset), if listed; b) browse the NHS Digital CTV3 to SNOMED Mapping Lookup ^8^ for relevant keywords and children. Find its SNOMED equivalent, if listed; c) Browse local TPP codes ^9^; d) Pragmatically browse the literature, online resources and white publications for further code indications ^9–13^; e) obtain clinical/programme input via the NHS England Digital First Primary Care programme (DFPC) on initially found codes of interest (long list), as well as further codes, to arrive at a refined list.

#### Box 2

Considerations when interrogating codes and coding activity

When interpreting output charts and tables it is important to consider that:

- All occurrences of codes are included and they do not necessarily indicate unique or new events (e.g. one patient encounter could generate several similar codes, one patient might have similar diagnoses recorded multiple times over time, or practices might bulk-import information).
- There might be other similar codes occurring in the data that are not included in the charts.
- Conversely, some codes are not exclusively used for the activity under study, e.g. remote consultations can include a broader range of activity such as telephone or video consultations.
- Not all codes represent activity occurring in general practice and may have been passed into the patient record from other services, including third party systems.
- Some apparent changes may represent changes in coding behaviour or displaced activities.
- Coding is dependent on manual input and therefore prone to inconsistency and gaps.

Coding related to online consultation systems and interpretation of recorded activity is not straightforward. This is due to a series of reasons:

- The use of online consultation systems and its national roll-out across practices are fairly recent. These codes do not differentiate between requests made using an online consultation system (route of access) and written online consultation appointments (mode of consultation with a patient/carer). These codes do not differentiate between administrative and clinical activity;
- There are not yet specific SNOMED codes for online consultation submissions and written online consultation appointments, although these are currently in development. In this analysis, coding (and clinical coding system) depends on the practice user, functionality and user interface within the GP IT clinical system, specific supplier technology and its template implementation;
- The implementation of online consultation systems can differ among practices, both in terms of the patient journey (service model) and the underlying technology. Some practices manage online requests and subsequent appointments within their OCS rather than GP IT clinical system. Therefore, the recording and nature of the (series of) codes generated will differ;
- The mode of making contact does not determine the mode of consultation, practices may only code the mode of consultation (appointment) e.g. a telephone, video or face to face appointment rather the route of contact; Where new codes have been created recently that are relevant to remote consultation or the use of online consultation systems, these are typically SNOMED and will tend to not have a CTV3 equivalent unless a local code is defined. Nevertheless, TPP is still quite reliant historically on CTV3 so the richness of recording will largely be in the legacy system (if using SNOMED, it may only do so via mapping to CTV3);
- In some cases, guidance is given for new forms of consultations to be recorded in annotated free-text fields of higher-level codes. Free text querying functionality is not currently available in OpenSAFELY.

### Recorded code activity and practice coverage over time [TPP]

The coding activity over time was characterised by individual code. The inter-practice variation was also assessed.

#### Cohort

Using OpenSAFELY-TPP, for each week or month (period) of study, the population of interest was defined as those aged one and over, alive and registered at the start of that period. Patients are assigned to the practice they are registered with in that period. In turn, any activity (OC-relevant codes, GP consultations) a patient has in that period is assigned to their practice of registration. This yielded a study population of over 23 million patients, relating to 2551 TPP practices.

#### Analysis

We extract the number of times each code was recorded a) over the period from January 2019 to December 2020 at monthly intervals and b) from the period of 6th January 2020 to the 22nd March 2021 at weekly intervals. The OpenSAFELY weekly data was specifically generated for contextualisation and benchmarking with a relevant weekly measure of online consultation submissions from a separate data source – the national OC/VC supplier collection^4^.

The OpenSAFELY monthly data was used for all further analysis. Absolute count instances and rates per 1,000 registered practice patient population were computed. The general practice consultation activity over that same period was also recorded for context - this uses a purpose-built function on OpenSAFELY rather than relying solely on counting code instances ^13 5^. Practice coverage, the number of practices with at least one instance of the code over the two year period, was also calculated, at both national and regional level ^14^. We also calculated the rate at which certain codes were recorded per 1,000 registered patients at a general practice level (among practices with any instance over the two year period) following methods described in [1]. We computed the deciles, median and interdecile range (IDR) for February, April, September and December 2020 (i.e. quarterly and for the start of the pandemic period).

### Sociodemographics of those with relevant coding activity [TPP]

Sociodemographic characteristics were characterised for those with any OC relevant coding instance.

#### Cohort

The population cohort was defined as all those registered with a single TPP GP practice between January 2019 and December 2020, resulting in a cohort of 20 million patients. The following characteristics were recorded, typically based on January 2019 status: ethnicity (based on ethnicity codelists); Sex; Age; Care home status ^15^; Household size; Practice registered with and associated region; Rurality of place of residence; Disability status (learning disabilities and intellectual disabilities codelists created from the QOF register); Deprivation quintile.

#### Analysis

When producing summary statistics, the study population was divided into whether the patients had had any recorded “online consultation”-relevant instance or not (at least one match for any of the shortlisted OC codes in the January 2019-December 2020 period).

Summary statistics were also computed for patients that had had any GP consultation in that same period. For a given sociodemographic or geographic dimension at a time, we computed for the two-year period:

- The instance rate of codes (number of code instances, standardised per 1,000 registered practice patient population)
- The coverage rate of codes (portion of population with at least one code instance)

### Clinical history of those with eConsultation activity [TPP]

In further follow-up exploratory analysis, the clinical history of patients with eConsultation activity was investigated.

#### Cohort

TPP practice registered patients with a single practice between 1^st^ March 2019 and 28^th^ February 2021. Occurrences of the eConsultation code or of GP-patient interaction were recorded. Only patients in practices with non-nil eConsultation coverage were considered. Age and gender were captured. Clinical history flags per patient were dictated by whether each patient had any recorded occurrence from the individual codelists prior to March 2019. These were chosen to broadly align with the most prevalent long-term conditions according to the NHSE Population and Person Insight dashboard and framework ^16^. The codelists were: hypertension, asthma, chronic respiratory disease other than asthma, osteoarthritis, depression, diabetes, chronic heart disease, cancer, atrial fibrillation, stroke, peripheral arterial disease, heart failure, chronic kidney disease, serious mental illness.

#### Analysis

Results, in terms of historic prevalence of individual clinical conditions among those with an eConsultation code in either the “pre-pandemic” (March 2019-February 2020) or “pandemic” (March 2020-February 2021) period were captured. Prevalence figures were also tabulated alongside those of two relevant comparator sub-cohorts (only within practices using the eConsultation code at all in that period) a) all the remaining practice population; b) the remaining practice population with a GP consultation recorded activity in that period. A multivariate logistic regression model was employed to assess which clinical conditions were associated with higher adjusted odds of having had an eConsultation code recorded, among the population in those practices that had had a GP consultation or eConsultation coding activity. Age groups and gender were also included as a first order case-mix adjustment. Further confounding (whether from interactions or other omitted characteristics) may remain.

### Recorded code activity and practice coverage [EMIS and TPP]

For the analysis of coding activity leveraging both EMIS and TPP (circa 99% of GP practices), a more fixed-scope exploration was implemented as a compromise given the OpenSAFELY-EMIS functionality and server availability, which was in its earlier stages. Results can be found in the Supplementary Information Appendix 4.

#### Cohort

We defined the population of interest as those aged one and over, alive, and registered as of 1 January 2019, leading to over 23 million and 30 million patients for TPP and EMIS respectively. Patients were assigned to the practice they were registered with at the start. In turn, any activity (codes associated with online consultation systems, remote consultations or with the EHR query on GP consultations) a patient had in that 2-year period was assigned to the initial practice of registration, rather than having activity reassigned to a new practice if the patient moved – this was necessary due to practical OpenSAFELY-EMIS functionality considerations. This differs from the TPP coverage analysis in previous sections, which did reflect month-on-month fluctuations in patient registration for each patient and reassigned activity accordingly.

#### Analysis

A table comparing EMIS and TPP statistics under similar conditions is presented. We show the number of times each code was recorded over the period from January 2019 to December 2020, aggregated over the 2 years. Values are given per 1,000 registered population and also per 1,000 registered patient population in practices where the code was recorded. To mitigate practice disclosure, results are not shown where a code was found in fewer than 5 practices.

### Software and Reproducibility

Data management was performed using Python 3.8, with analysis carried out using R. All code used for data management and analysis is shared openly for review and re-use under an MIT open license at https://github.com/opensafely/OS_OC_v001-research. The developed codelist can be found at https://www.opencodelists.org/codelist/user/martinaf/online-consultations-snomed-v01/28bba9bc/.

### Patient and Public Involvement

We have developed a publicly available website https://opensafely.org/ through which we invite any patient or member of the public to contact us regarding this study or the broader OpenSAFELY project.

## Results

Results and commentary are given below for TPP based practices and cohorts. Results for the combined OpenSAFELY-TPP and OpenSAFELY-EMIS analysis can be found in the Supplementary Information.

### Weekly coding activity and contextualisation with rapid supplier collection

The top graph in Figure 1 depicts the absolute instances of code eConsultation, as well as of all the SNOMED codes together. It reflects coding activity through all TPP practices in the 6th January 2020-22nd March 2021 period. Different practices may approach the coding of different activity associated with the use of online consultation systems differently though, especially based on the OC supplier system in place, chosen OC pathways and implementation maturity ^17^.

**Figure 1.**
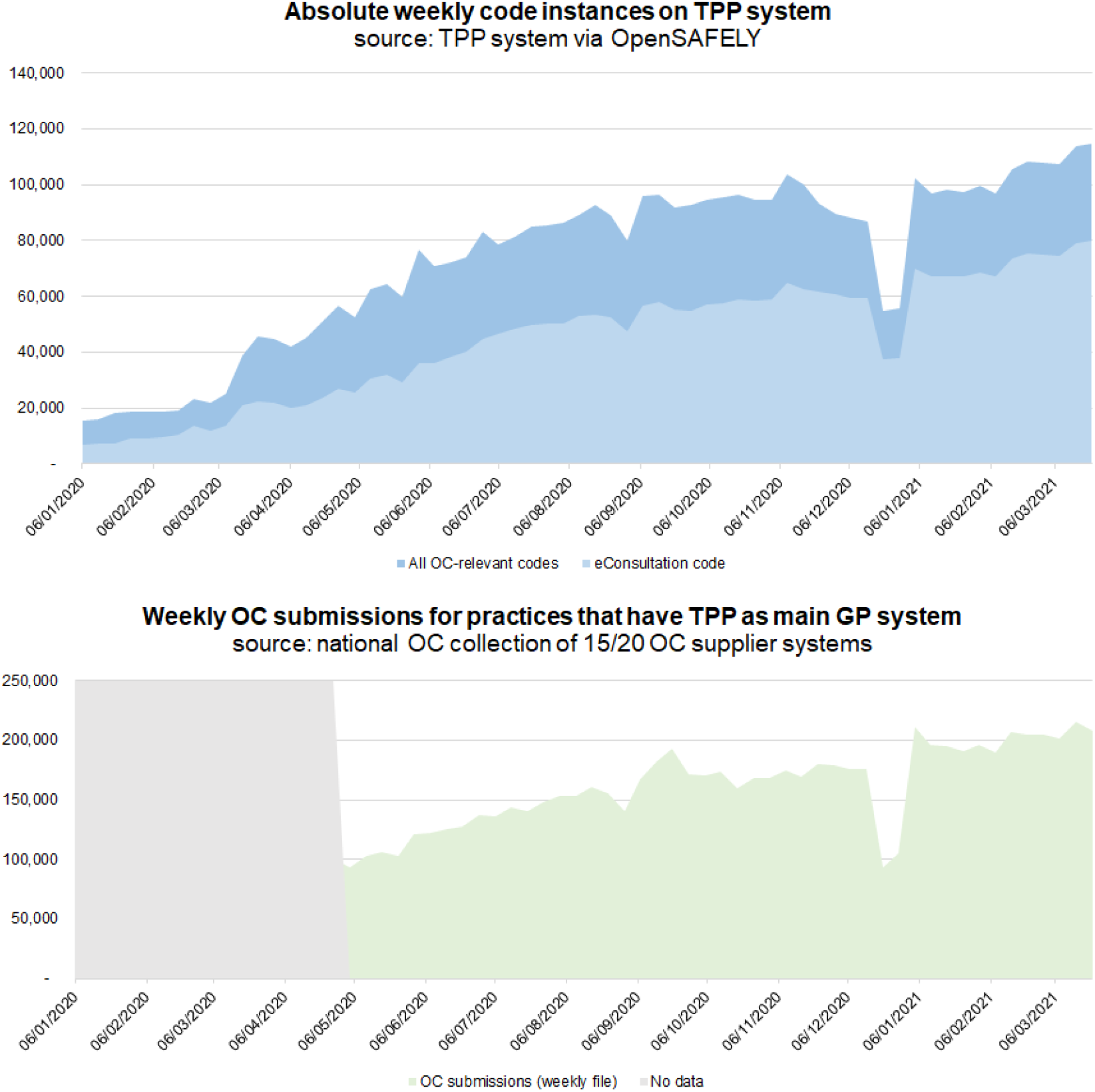
Top: absolute weekly online consultation code instances in TPP System (source: OpenSAFELY-TPP). Bottom: weekly OC submissions for practices that have TPP as main GP system (source: national rapid collection. TPP practices were identified via the POMI data collection^6^. Between 66-132 practices each week had no clear system associated in POMI and were not included.

Separately, the bottom graph in Figure 1 depicts the total online consultation submissions in the period of weeks commencing 27th April 2020-22nd March 2021 according to the NHS England OC/VC data collection from system suppliers (rapid and aggregate) ^4^. This is likely still an underestimation due to data completeness.

The two data sources track different but related activity. The shape of the graphs over time looks the same in terms of peaks and troughs. If we assume each OC submission should generate at least one activity code in primary care systems, then the data suggests that coding activity in GP IT clinical systems is not fully tracking all OC activity. This may, however, relate to a range of reasons including: codes such as eConsultation only being triggered downstream from what is considered a submission in an OC system; certain practices or OC systems not yet using dedicated codes; certain practices or OC systems using codes that are different or broader than those studied here; how an OC submission is defined within the supplier collection; practices using some codes for clinical activity but not administrative; certain practices potentially not recording all OC activity in the GP IT clinical system, for example repeat requests or some practices managing online requests and subsequent appointments within their online consultation system rather than GP IT clinical system.

### Codes in use and practice coverage

Figure 2 shows the portion of practices that had at least one instance of the respective SNOMED code, over the two year period. Breakdowns by region are given in Appendix 1.

**Figure 2.**
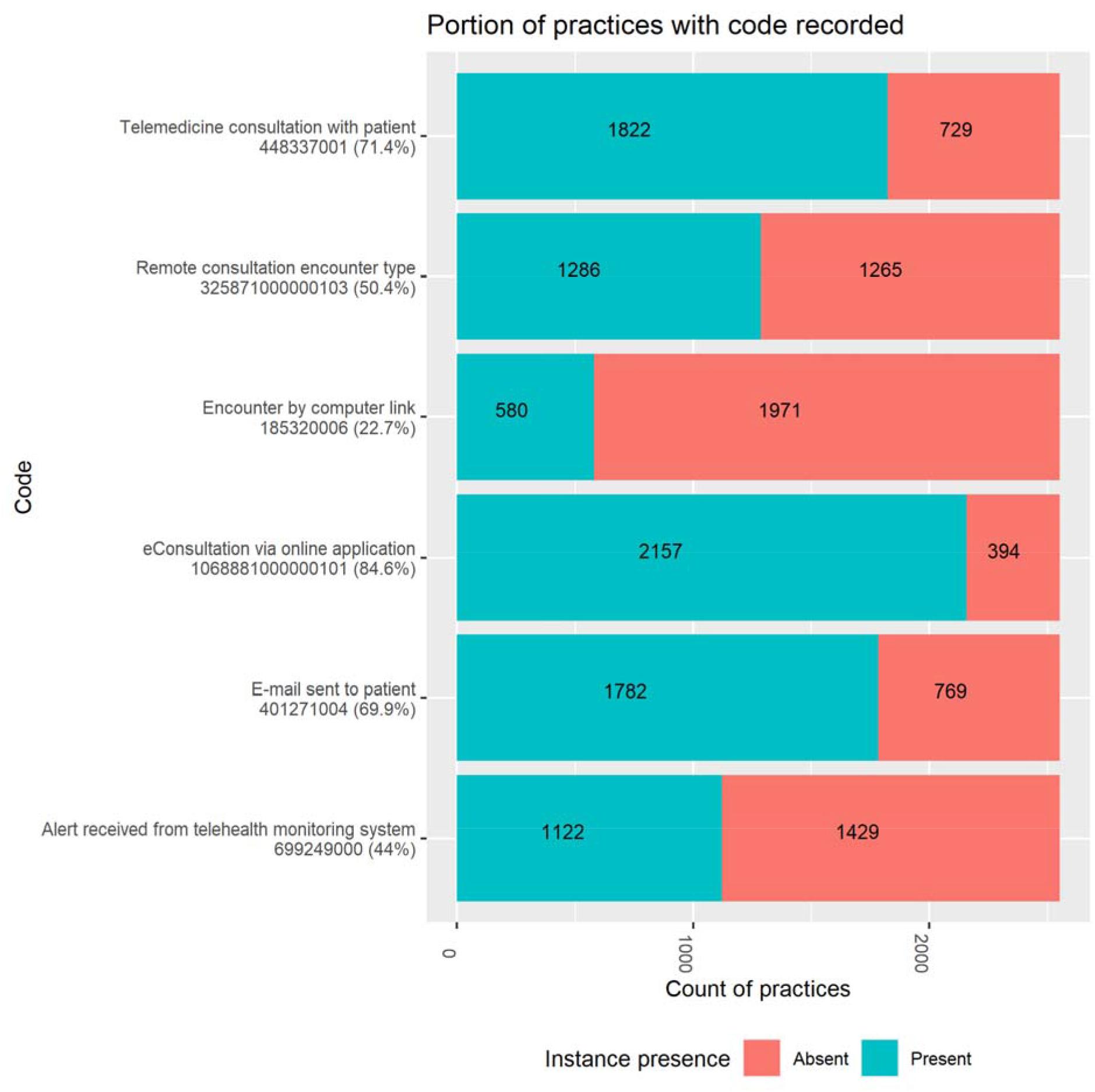
Portion of TPP practices with any recorded activity for online consultation relevant codes in general practice (January 2019 - December 2020). Codes with no activity at all omitted.

12 of 18 codes returned no results in TPP. The last column of Table 1 indicates this. The SNOMED codes for which instances *were* found also correspond to those where both: a) a CTV3 mapping was available when specifying the codelists; b) CTV3-querying had activity recorded (not shown). The practice coverage was, in decreasing order: eConsultation via online application (1068881000000101) - 85% of practices; Telemedicine consultation with patient (448337001) - 71% of practices; E-mail sent to patient (401271004) - 70% of practices; Remote consultation encounter type (325871000000103) - 50% of practices; Alert received from telehealth monitoring system (699249000) - 44% of practices; Encounter by computer link (185320006) - 23% of practices.

### Monthly trends in coding activity

While the previous plots focussed on coverage, Figure 3 shows the monthly utilisation of the various codes (coding activity) over the period from January 2019 to December 2020. Values are given as a rate (per 1,000 cohort population). The entire cohort population is considered, rather than just those in practices where each code has been recorded. The rate of GP consultation events is also given, for context (its practice coverage is near complete at over 99%, as expected).

**Figure 3.**
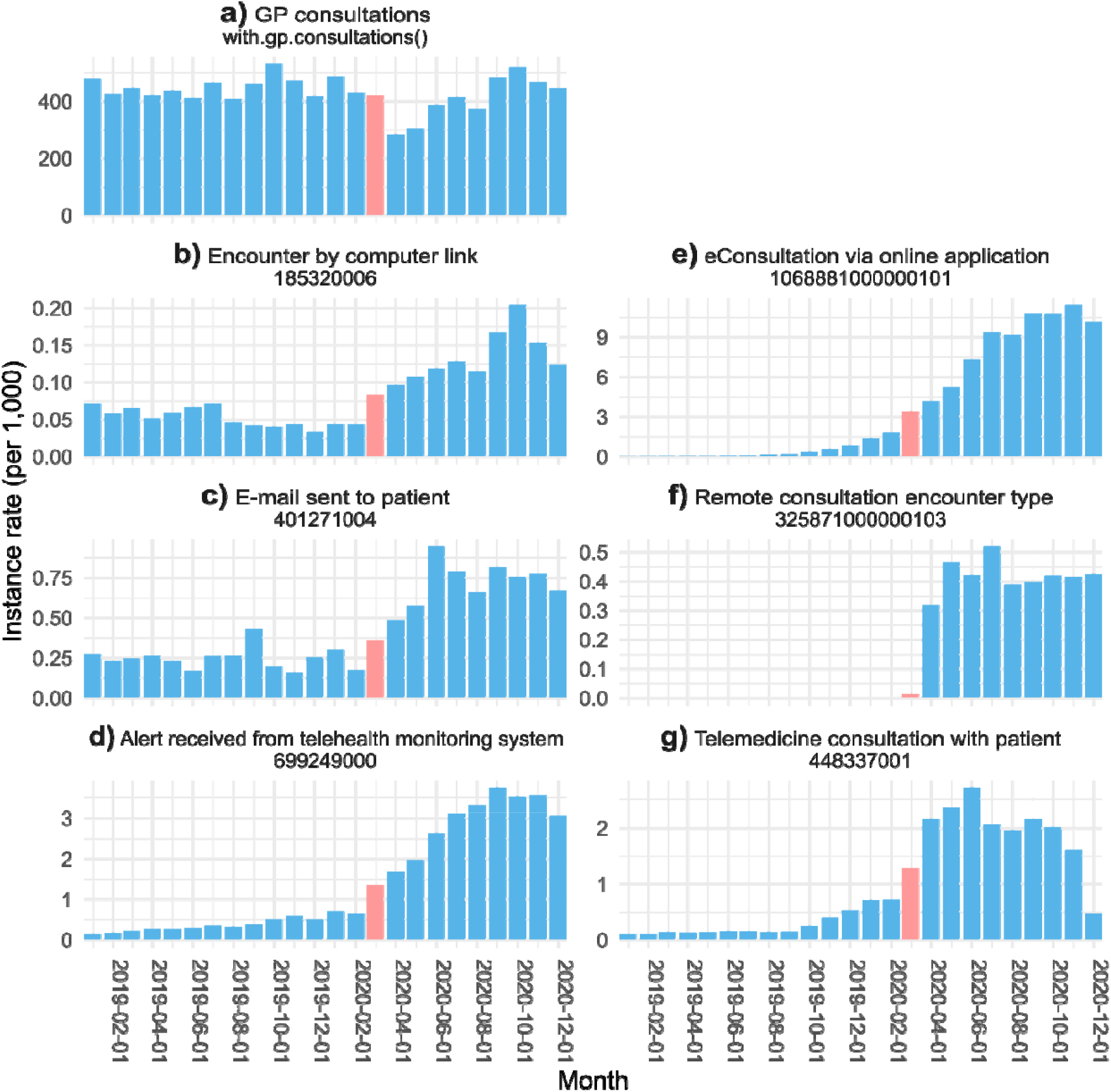
Monthly code instance rates per 1,000 registered practice patient population of SNOMED codes in TPP general practice (January 2019 - December 2020). March 2020 indicated in pink. Figure with absolute counts given in Appendix 1.

The codes with highest activity were, in order of highest monthly peak:

- eConsultation via online application (1068881000000101) - a peak of over 10 monthly coding events per 1,000 registered population in November 2020. This has increased rapidly from virtually none in early 2019 (Figure 3e).
- Alert received from telehealth monitoring system (699249000) - a peak of over 3.5 events per 1,000 registered population in September 2020. This has increased rapidly compared to 2019. A first step-change is seen around the start of the pandemic (from February to March 2020) (Figure 3d).
- Telemedicine consultation with patient (448337001) - a peak of over 2.5 events per 1,000 registered population in June 2020. Step changes from February to March and March to April 2020 are noticeable (Figure 3g).
- E-mail sent to patient (401271004) - a peak of close to 1 event per 1,000 registered population in June 2020. Step changes from February to March and March to April 2020 are noticeable (Figure 3c).
- Remote consultation encounter type (325871000000103) - a peak of over 0.5 events per 1,000 registered population in July 2020. This is likely a new code - its use appears to be first recorded in March 2020. This may relate to TPP introducing a local-TPP ^18^ dedicated code that maps to this SNOMED code (Y22b4) (Figure 3f).
- Encounter by computer link (185320006) - a peak of over 0.2 events per 1,000 in October 2020. Its use seemed to be in slight decline in 2019 and then got a step increase from March 2020 (Figure 3b).
- We have also plotted the monthly rate of (overall) GP consultations regardless of the modality in the TPP practices. This stood broadly above 400 consultations per 1,000 patients in 2019. The dip is seen around April 2020. Recovery occurred, with October 2020 registering the second highest monthly rate (519 consultations per 1,000 patients), after October 2019 (532 consultations per 1,000 patients) (Figure 3a).

### Inter-practice variation in monthly coding activity trends

To better convey the coding activity over time and in terms of inter-practice variation, the median and decile trends were created: ‘eConsultation’ (Figure 5), ‘telemedicine consultation with patient’ (Figure 6), aggregate of all codes (Figure 7), and contextual highlighting GP consultations (Figure 4). Deciles illustrate variation across practices for a given metric in a more compact form; for each timepoint, practices are sorted and ranked from lowest to highest activity, with points that define the top 10%, 20%, …, 80%, 90% respectively plotted for each month. The 50% decile, i.e. the median, is shown as a continuous, bold line. Figure 7 shows that there is considerable variation in coding activity levels across practices, with the top deciles of practices gaining much of their activity around the start of the pandemic.

**Figure 4.**
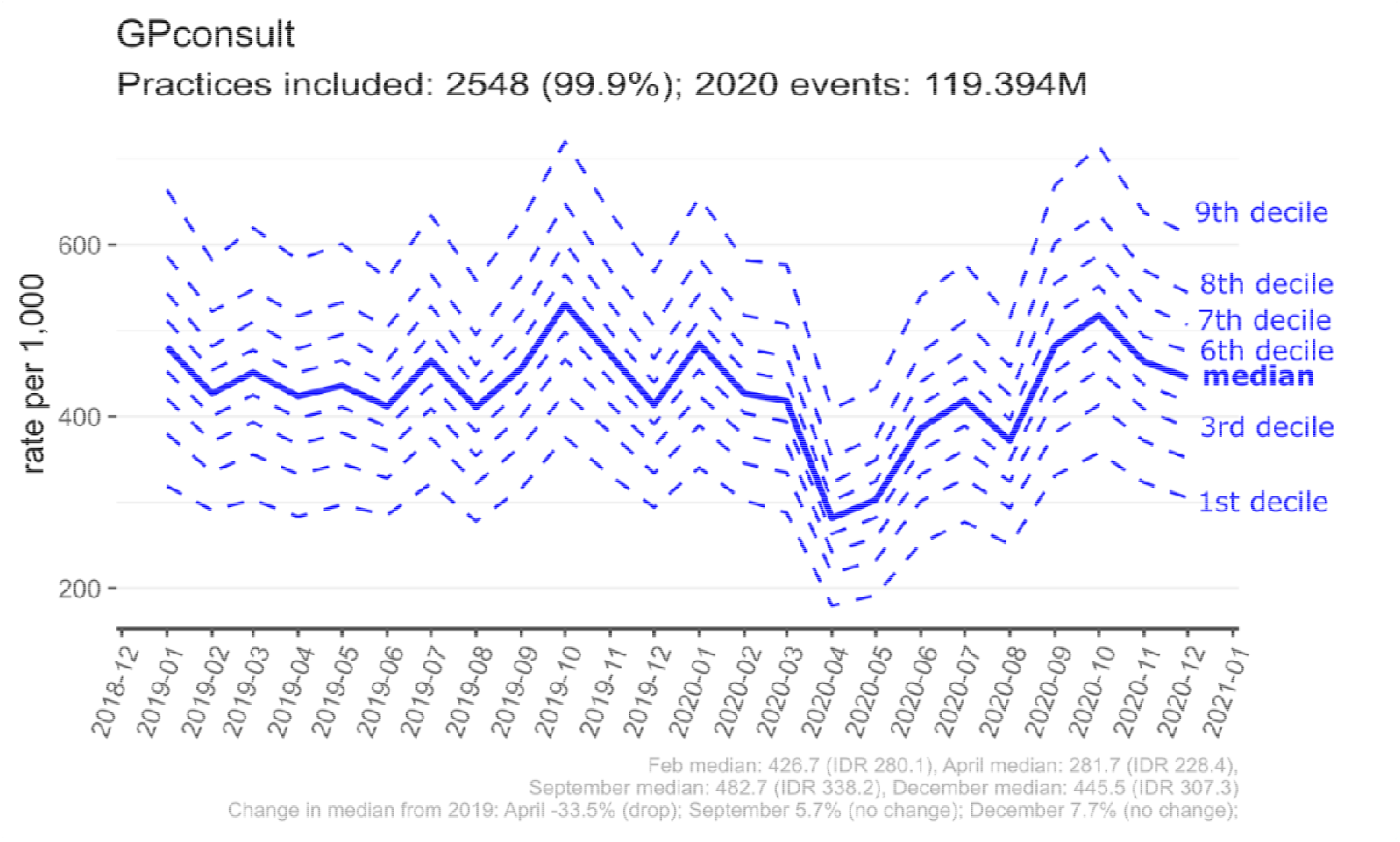
Contextual - Recording of results from GP Consultations (any modality) in general practice (January 2019 - December 2020)

**Figure 5.**
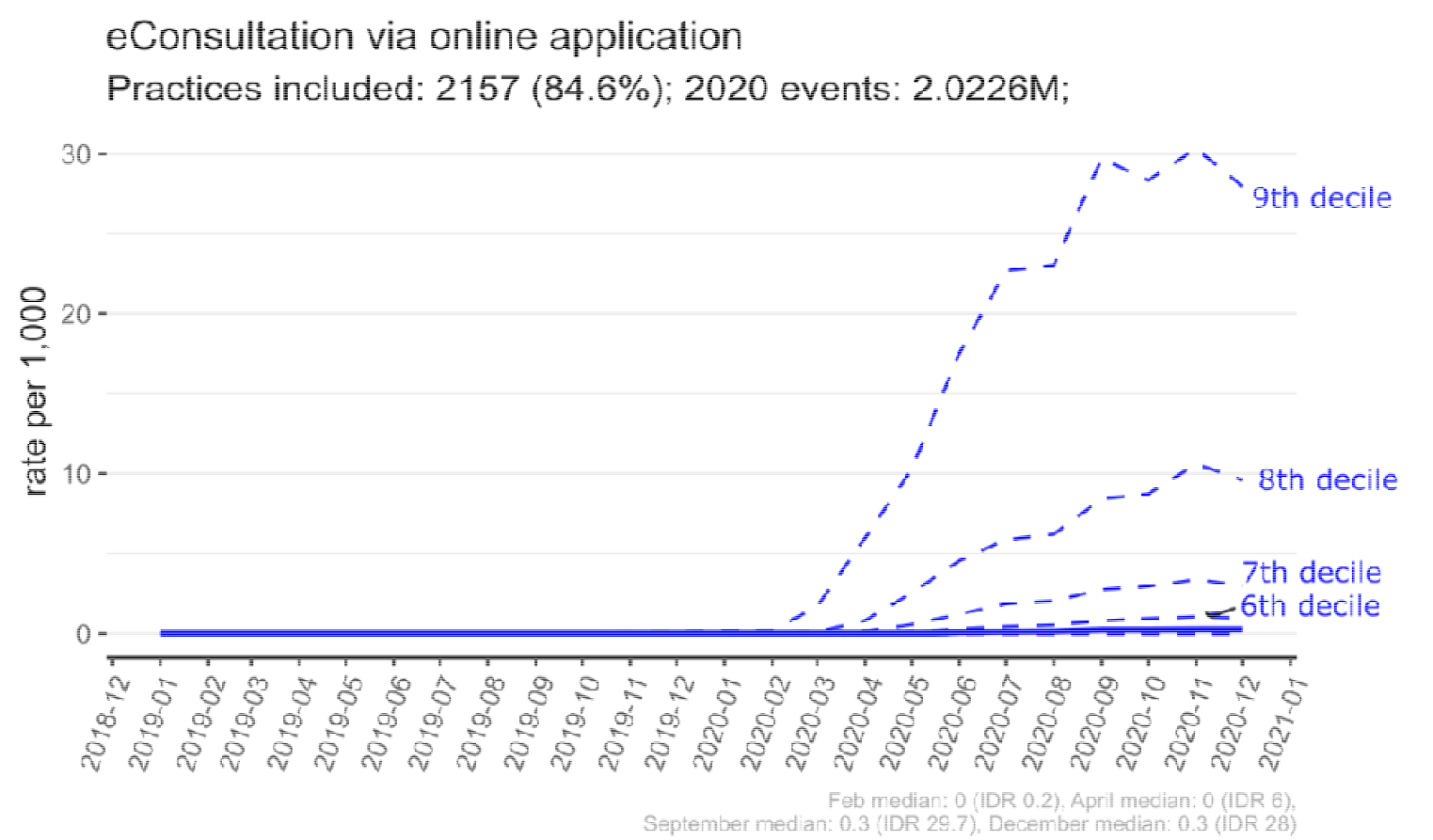
Recording of results from eConsultation via Online Application (“1068881000000101” ∼ “Y1f3b”) in general practice (January 2019 - December 2020). The top four deciles can be discerned, with the top practice decile peaking around 30 events per 1,000 patients. The lowest four deciles of practices have very low rates, hence why they cannot be easily discerned.

**Figure 6.**
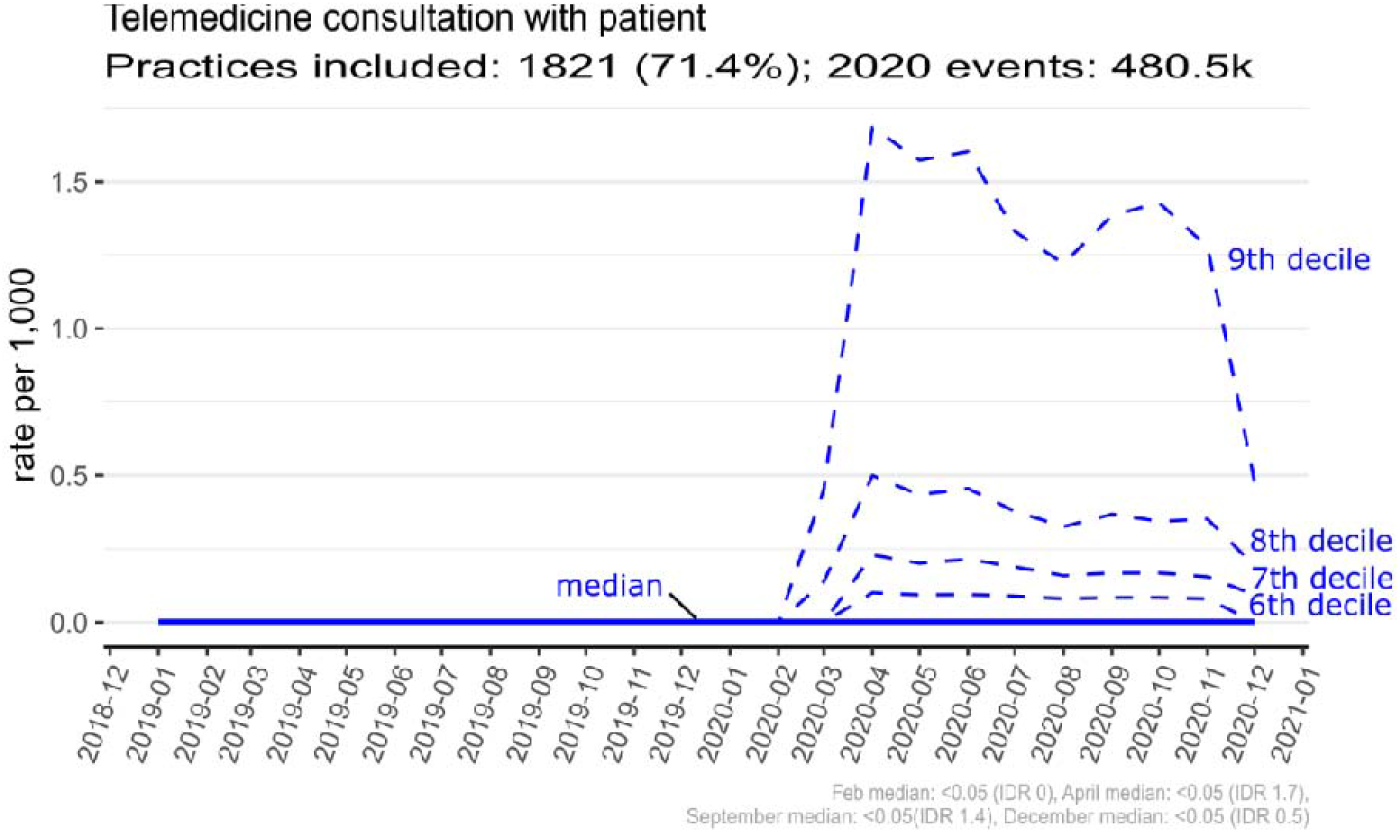
Recording of results from Telemedicine consultation with patient (“448337001” ∼ “XaXcK”) in general practice (January 2019 - December 2020). The top four deciles can be discerned, with the top practice decile peaking above 1.5 events per 1,000 patients. The lowest four deciles of practices have very low rates, hence why they cannot be easily discerned.

**Figure 7.**
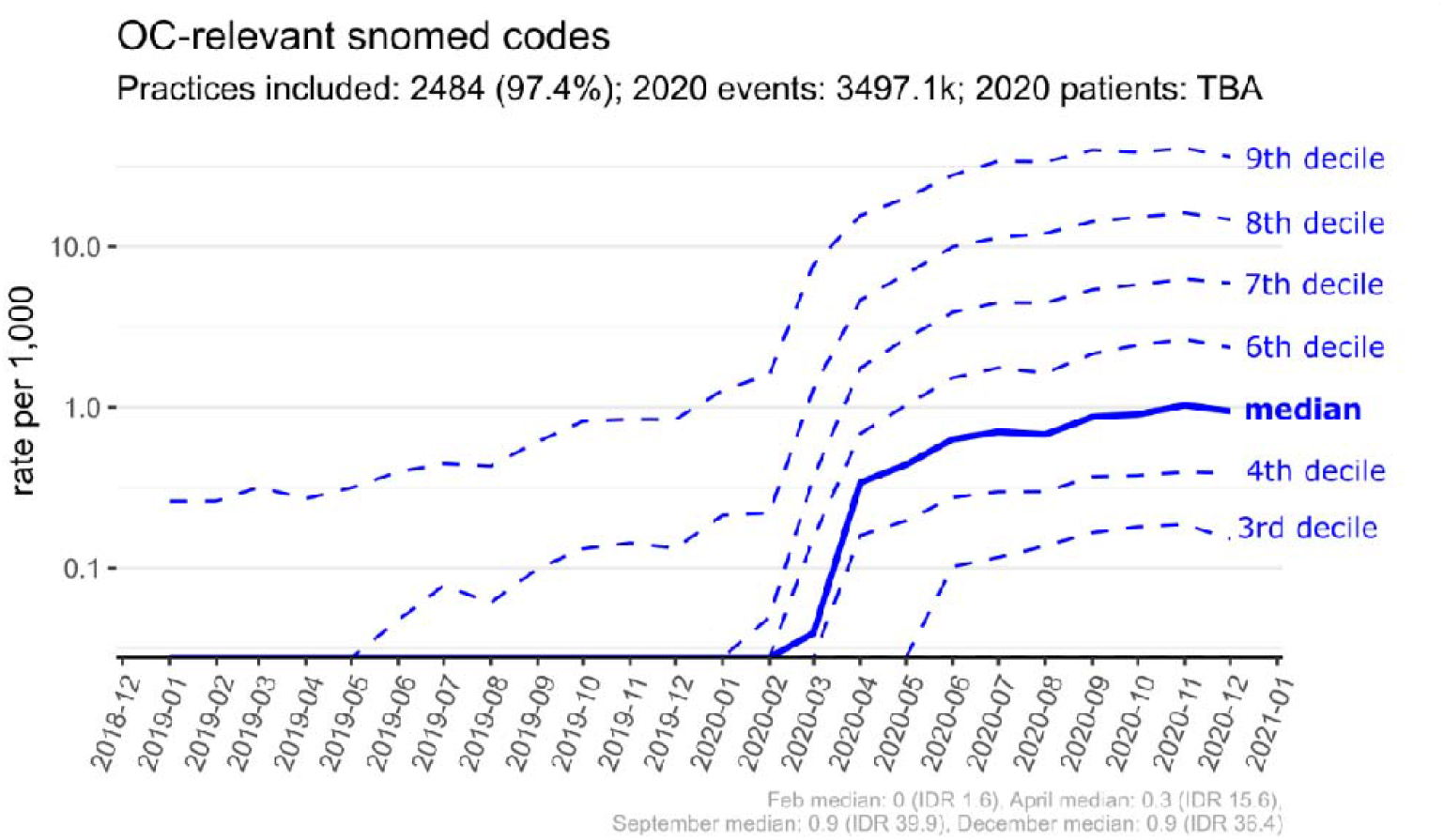
Recording of results from any of the shortlisted SNOMED online consultations codes in general practice (January 2019 - December 2020). Log Scale. The top practice decile peaks above 36 events per 1,000 patients.

### Demographic patterns in online consultation coding activity

Broadly, Table 2 shows that the cohort with at least one “online consultation”-relevant coding instance has a higher preponderance of female patients; has a higher relative preponderance of those aged 18-40, followed by those aged 40-50 and 50-60; skews more towards white patients; skews more towards those who are least deprived.

**Table 2.** Characteristics of the studied cohort, both overall and by a) patients without a recorded online consultation related code instance; b) patients with such an instance. The p-value indicates significance for the difference in distributions at the 99.9%CI

| Characteristic | Overall,<br>N = 20,651,036<sup>1</sup> | Had any OC-relevant coding instance |  | p-value<sup>2</sup> |
| --- | --- | --- | --- | --- |
|  |  | NO, N = 19,563,117<sup>1</sup> | YES, N = 1,087,919<sup>1</sup> |  |
| **sex** |  |  |  | <0.001 |
| Female | 10,260,731 (50%) | 9,599,496 (49%) | 661,235 (61%) |  |
| Male | 10,260,731 (50%) | 9,963,322 (51%) | 426,654 (39%) |  |
| Other/Unknown | 329 (<0.1%) | 299 (<0.1%) | 30 (<0.1%) |  |
| **age** | 41 (22, 59) | 41 (21, 59) | 43 (27, 58) | <0.001 |
| **Age group** |  |  |  | <0.001 |
| (0,18] | 4,298,691 (21%) | 4,151,378 (21%) | 147,313 (14%) |  |
| (18,40] | 5,738,142 (28%) | 5,388,980 (28%) | 349,162 (32%) |  |
| (40,50] | 2,842,130 (14%) | 2,665,869 (14%) | 176,261 (16%) |  |
| (50,60] | 2,913,528 (14%) | 2,735,067 (14%) | 178,461 (17%) |  |
| (60,70] | 2,269,212 (11%) | 2,144,742 (11%) | 124,470 (12%) |  |
| (70,80] | 1,673,588 (8.2%) | 1,598,702 (8.2%) | 74,886 (6.9%) |  |
| (80,Inf] | 746,742 (3.6%) | 716,706 (3.7%) | 30,036 (2.8%) |  |
| Unknown | 169,003 | 161,673 | 7,330 |  |
| **ethnicity** |  |  |  | <0.001 |
| Asian | 1,252,414 (6.1%) | 1,209,218 (6.2%) | 43,196 (4.0%) |  |
| Black | 412,399 (2.0%) | 398,242 (2.0%) | 14,157 (1.3%) |  |
| Mixed | 249,470 (1.2%) | 238,762 (1.2%) | 10,708 (1.0%) |  |
| Other | 6,026,577 (29%) | 5,737,108 (29%) | 289,469 (27%) |  |
| White | 12,710,176 (62%) | 11,979,787 (61%) | 730,389 (67%) |  |
| **living alone** | 5,783,003 (28%) | 5,466,461 (28%) | 316,542 (29%) | <0.001 |
| **region** |  |  |  | <0.001 |
| East | 4,823,404 (23%) | 4,623,066 (24%) | 200,338 (18%) |  |
| East Midlands | 3,618,902 (18%) | 3,458,477 (18%) | 160,425 (15%) |  |
| London | 1,340,024 (6.5%) | 1,277,438 (6.5%) | 62,586 (5.8%) |  |
| North East | 963,807 (4.7%) | 960,313 (4.9%) | 3,494 (0.3%) |  |
| North West | 1,843,088 (8.9%) | 1,722,626 (8.8%) | 120,462 (11%) |  |
| South East | 1,357,871 (6.6%) | 1,236,531 (6.3%) | 121,340 (11%) |  |
| South West | 2,838,383 (14%) | 2,586,842 (13%) | 251,541 (23%) |  |
| West Midlands | 861,670 (4.2%) | 840,558 (4.3%) | 21,112 (1.9%) |  |
| Yorkshire & The Humber | 2,997,813 (15%) | 2,851,255 (15%) | 146,558 (13%) |  |
| Unknown | 6,074 | 6,011 | 63 |  |
| **deprivation quintile** |  |  |  | <0.001 |
| Q1 (most) | 4,157,772 (20%) | 3,989,883 (21%) | 167,889 (16%) |  |
| Q2 | 4,032,329 (20%) | 3,822,954 (20%) | 209,375 (20%) |  |
| Q3 | 4,259,619 (21%) | 4,023,228 (21%) | 236,391 (22%) |  |
| Q4 | 4,052,737 (20%) | 3,817,032 (20%) | 235,705 (22%) |  |
| Q5 (least) | 3,796,821 (19%) | 3,577,294 (19%) | 219,527 (21%) |  |
| Unknown | 351,758 | 332,726 | 19,032 |  |
| **rural urban** |  |  |  | <0.001 |
| Other | 328,860 (1.6%) | 310,706 (1.6%) | 18,154 (1.7%) |  |
| Rural | 4,113,110 (19.9%) | 3,896,532 (19.9%) | 216,578 (19.9%) |  |
| Urban | 16,209,066 (78.4%) | 15,355,879 (78.4%) | 853,187 (78.4%) |  |
| **care home** |  |  |  | <0.001 |
| Yes | 37,137 (0.2%) | 34,545 (0.2%) | 2,592 (0.2%) |  |
| Non | 20,613,899 (100%) | 19,528,572 (100%) | 1,085,327 (100%) |  |

<sup>1</sup> n (%); Median (IQR)
<sup>2</sup> Pearson's Chi-squared test; Wilcoxon rank sum test

Patterns by gender and age are broadly in line with those found in a previous pilot in the South West, using ‘eConsult’ ^10^. The results may also be reflective of the greater implementation challenges such as time, capacity and support required to embed the use of digital tools in practices working in the most challenging circumstances and highest areas of deprivation, communications and language to support people to navigate access points and general practice involvement in local commissioning decisions which were made at pace due to the urgency of responding to the pandemic. Alongside support with patient factors such as health and digital literacy, confidence with and access to digital devices and data. Furthermore, there is variation in the design, functionality and interoperability of online consultation systems which may impact the usability and accessibility of different systems, this study does not explore rates of use across different demographic characteristics between different types of online consultation system.

For further contextualisation of results in Table 2, the distribution of the full study population based on whether they had general practice consultations (as opposed to not) is given in Appendix 2. The differential patterns between those with an OCS instance and those with wider general practice interaction are similar to those found when comparing it with the full cohort population in Table 2, albeit in a less pronounced way, both in rates and coverage. Namely, we still observe higher relative preponderance of those aged 18-40 and higher preponderance among those least deprived, non-BAME or female.

Further breakdowns per sociodemographic characteristic, coding coverage and instance rates are also given in the Appendix 2. While instances of codes associated with online consultation systems or remote online consultations cannot be compared directly to GP consultation figures or GPAD figures, it is useful to look at relative values across levels of a given sociodemographic characteristic. Some sub-cohorts are small so figures and differential patterns require caution given underlying uncertainty.

### Patterns in clinical history for those with ‘eConsultation’ coding activity

The cohort of patients that in March 2020 - February 2021 had eConsultation activity coded in their records was characterised overall by a lower prevalence (clinical history) of most long-term conditions compared to the remaining population with GP-consultation recorded activity that year (Table 3). Notable exceptions were asthma and depression, where respectively 20% and 23% of eConsultation patients had a clinical history of these, against 17% and 19% for other patients with general GP-patient interactions. The comparison against the full population in those practices (rather than just patients therein with GP-consultation recorded activity) is given in Appendix 3 (Table A8), producing a more stark difference for asthma and depression but a more homogenous profile otherwise (indicating that online consultation patients resemble more the general population, clinical history-wise). The tabulation for the “pre-pandemic” eConsultation activity is also given Appendix 3 (Table A9).

**Table 3.** Clinical history characteristics of the cohort with eConsultation code recorded in March 2020-February 2021. Comparison against population in those practices that had GP consultation recorded.

| Clinical history (pre-March 2019)# | Overall, N = 9,835,747<sup>1</sup> | Had eConsultation code instance in Mar20-Feb21 (among those with an eConsult/GP consultation) |  | p-value<sup>2</sup> |
| --- | --- | --- | --- | --- |
|  |  | No, N = 9,018,200<sup>1</sup> | Yes, N = 817,547<sup>1</sup> |  |
| history_hypertension | 2,166,059 (22%) | 2,035,381 (23%) | 130,678 (16%) | <0.001 |
| history_asthma | 1,716,492 (17%) | 1,551,624 (17%) | 164,868 (**20%**) | <0.001 |
| history_osteoarthritis | 1,519,293 (15%) | 1,426,313 (16%) | 92,980 (11%) | <0.001 |
| history_depression | 1,941,426 (20%) | 1,754,903 (19%) | 186,523 (**23%**) | <0.001 |
| history_diabetes | 996,641 (10%) | 938,575 (10%) | 58,066 (7.1%) | <0.001 |
| history_chronic_heart_disease | 617,213 (6.3%) | 583,834 (6.5%) | 33,379 (4.1%) | <0.001 |
| history_cancer | 531,640 (5.4%) | 497,628 (5.5%) | 34,012 (4.2%) | <0.001 |
| history_atrial_fibrillation | 282,954 (2.9%) | 268,036 (3.0%) | 14,918 (1.8%) | <0.001 |
| history_stroke | 197,873 (2.0%) | 188,013 (2.1%) | 9,860 (1.2%) | <0.001 |
| history_chronic_respiratory_disease | 404,974 (4.1%) | 381,487 (4.2%) | 23,487 (2.9%) | <0.001 |
| history_peripheral_arterial_disease | 89,347 (0.9%) | 85,317 (0.9%) | 4,030 (0.5%) | <0.001 |
| history_heart_failure | 152,507 (1.6%) | 144,499 (1.6%) | 8,008 (1.0%) | <0.001 |
| history_chronic_kidney_disease | 13,393 (0.1%) | 12,426 (0.1%) | 967 (0.1%) | <0.001 |
| history_serious_mental_illness | 114,139 (1.2%) | 106,557 (1.2%) | 7,582 (0.9%) | <0.001 |
| Econsult_pre_had<sup>3</sup> | 53,521 (0.5%) | 15,335 (0.2%) | 38,186 (4.7%) | <0.001 |
| Gp_consult_pre_had<sup>3</sup> | 8,541,965 (87%) | 7,829,178 (87%) | 712,787 (87%) | <0.001 |
| Gp_consult_post_had<sup>3</sup> | 9,811,243 (100%) | 9,018,200 (100%) | 793,043 (97%) | <0.001 |

<sup>1</sup> n (%)
<sup>2</sup> Pearson's Chi-squared test (univariate tests)
<sup>3</sup> 'Pre' refers to Mar19-Feb20. 'Post' refers to Mar20-Feb21

In part, this overall lower prevalence may reflect the inherent nature of the intended online consultation submissions themselves, which are not exclusively focused on the need for a traditional GP consultation but reflect more general population needs such as admin tasks, digitally-enabled routine checks and queries.

Despite the profiling above, it is important to note that the OC systems user profile may not be generalisable and may be very dependent on the practice-by-practice model. Namely, user profile may be influenced by how the online consultation system has been implemented, who and for what conditions practices have promoted the online consultation option to, the type of online consultation system, ease of finding and navigating the online consultation system and staff confidence in using digital tools. There may also be differences in user profile in practices with high utilisation of online consultation systems compared to practices with low utilisation. Authors of the 2016 South West pilot study did note that the practices involved in the early pilot had fewer patients with long-term health conditions than practices in the rest of England, reflecting potentially greater early-adopter appetite or capability by such practices.^10^

From the previous section, we have also seen that activity skews to a younger and female-tilted profile, which may be masking some of the clinical history profile characteristics. Table 4 shows that, when considering all clinical history conditions simultaneously, while also factoring in age and gender (first order adjustment), the clinical characteristics that were *more* prevalent among those with eConsultation activity recorded expanded to (odds ratio (OR) >1 and significant) :

**Table 4.** Adjusted odds of having had an online consultation in March 2020-February 2021 given past clinical history, age and gender. Odds ratio (OR) considered against the remaining population in those practices that had any GP consultation (GP-patient interaction) recorded during that period.

| Characteristic | Odds ratio | Pr(>\|z\|) | LCI95 | UCI95 s.s. |
| --- | --- | --- | --- | --- |
| (Intercept) | 0.058 | <0.001 | 0.057 | 0.058 * |
| sexMale | ref |  |  |  |
| sexFemale | 1.206 | <0.001 | 1.200 | 1.211 * |
| sexOther/Unknown | 1.490 | 0.068 | 0.971 | 2.288 |
| history_hypertension | 1.015 | <0.001 | 1.008 | 1.023 * |
| history_asthma | 1.131 | <0.001 | 1.124 | 1.137 * |
| history_osteoarthritis | 1.048 | <0.001 | 1.040 | 1.057 * |
| history_depression | 1.144 | <0.001 | 1.138 | 1.151 * |
| history_diabetes | 0.858 | <0.001 | 0.851 | 0.866 * |
| history_chronic_heart_disease | 0.965 | <0.001 | 0.953 | 0.977 * |
| history_cancer | 1.080 | <0.001 | 1.068 | 1.093 * |
| history_atrial_fibrillation | 1.119 | <0.001 | 1.099 | 1.139 * |
| history_stroke | 0.914 | <0.001 | 0.895 | 0.933 * |
| history_chronic_respiratory_disease | 0.928 | <0.001 | 0.915 | 0.941 * |
| history_peripheral_arterial_disease | 0.897 | <0.001 | 0.868 | 0.926 * |
| history_heart_failure | 1.043 | 0.001 | 1.018 | 1.069 * |
| history_chronic_kidney_disease | 0.967 | 0.312 | 0.905 | 1.033 |
| history_serious_mental_illness | 0.725 | <0.001 | 0.708 | 0.742 * |
| age_group(0,18] | 1.308 | <0.001 | 1.295 | 1.321 * |
| age_group(18,40] | 1.940 | <0.001 | 1.923 | 1.957 * |
| age_group(40,50] | 1.665 | <0.001 | 1.650 | 1.681 * |
| age_group(50,60] | 1.381 | <0.001 | 1.369 | 1.394 * |
| age_group(60,70] | ref |  |  |  |
| age_group(70,80] | 0.681 | <0.001 | 0.673 | 0.689 * |
| age_group 80+ | 0.533 | <0.001 | 0.524 | 0.542 * |

- Hypertension (OR: 1.015 [1.008-1.023])
- Asthma (OR: 1.131 [1.124-1.137])
- Osteoarthritis (OR: 1.048 [1.040-1.057])
- Depression (OR: 1.144 [1.138-1.151])
- Cancer (OR: 1.080 [1.068-1.093])
- Atrial fibrillation (OR: 1.119 [1.099-1.139])
- Heart failure (OR: 1.015 [1.018-1.069])

Other clinical history conditions were mainly *less* prevalent with these adjustments.

## Discussion

### Summary

Using OpenSAFELY-TPP and OpenSAFELY-EMIS, we were able to generate data on clinical coding activity relevant to online consultation systems and remote monitoring, across circa 99% of practices and over 53 million patient records. We observed large variation in coding instance rates among practices in England and between the two EHR supplier systems. For TPP practices (circa 40% of practices in England), we explored further the trends and variation in coding activity related to digital forms of interaction. Coding activity increased rapidly over the study period, with a marked increase and acceleration at the start of the pandemic and first lockdown. Further, we found population sub-cohorts – both in terms of sociodemographics and clinical history - against which the recorded instance rates were higher.

Above all, this work highlighted that more needs to be done to consolidate and harmonise activity definition (by route and mode) and coding practices associated with the use of online consultations systems in general practice.

### Findings in context and comparison of existing evidence

Six of the eighteen codes identified were in active use in TPP practices. The ones used by more practices were, in order, eConsultation via online application, Telemedicine consultation with patient and e-mail sent to patient - in 70% or more of practices. In the analysis extension to OpenSAFELY-EMIS, its practices also had registered activity for these codes, but their volumes were quite different, indicating likely differences in digital supplier systems, coding approaches or population served. Code ‘eConsultation via online application’ has been explicitly linked to online consultations/triages, namely by supplier eConsult ^19^ and North of England Commissioning Support in their SystemOne guidance ^20^. Code ‘Consultation via multimedia encounter type’, detected in EMIS, has been suggested for use in NHS England total triage guidance ^21^.

Coding activity related to digital forms of interaction picked up rapidly from the start of the COVID-19 pandemic and the first lockdown, from March 2020. In the second semester of 2020, over nine monthly eConsultation coding events per 1,000 registered population were registered, compared to less than one per 1,000 a year prior. This broad rising trend observed for OCS codes was consistent with the rising trend in weekly online consultation submissions as captured in the NHSE OC/VC supplier collection ^4^, including broadly in terms of more contained peaks and troughs.

Over 2019 and 2020 respectively, 227,429 and 3,323,333 OCS relevant codes were found in the TPP cohort. When contextualised with wider GP consultation/interaction coding, this corresponded to 1.8 OCS codes for every 1000 GP consultation codes in 2019 and 27.9 OCS codes for every 1000 GP consultation codes in 2020. Though direct interpretation of this and other publication sources is complicated due to lack of coding standardisation, interpretation and completeness, this relative level of OCS utilisation (either route of access or written consultation mode) is broadly in line with observations from other sources. GPAD (Appointments in General Practice) is the central statistics publication on appointments that have taken place in general practice, showing namely a record number of over 30M recorded appointments in November 2021. In terms of appointment mode, in 2019 and 2020 respectively, 5.6 and 4.6 in 1,000 appointments had been found to be recorded mode-wise as either video, videoconference or online (i.e. written consultation), though publication caveats related to how mode of consultation is recorded and defined, field completion and the effect of the pandemic warrant caution and could mean levels are understated and conflated ^22^. Much of the partial or total ‘triage’ activity is also not reflected in the GPAD statistics collected and published. Further on appointment mode, in the self-reported GP Patient Survey (GPPS) 2020 and 2021 editions - each mainly covering experiences respectively of the year prior -, 0.2% and 2.6% respectively of those that had booked an appointment said that they got an appointment to speak to someone online (e.g. video or written) ^23^.

Inter-practice variation was large, reflecting the recent nature of online consultation system implementation adoption and use: December 2020 saw the median practice have 0.9 recorded codes per 1,000 population, compared to about 36 for the highest decile of practices.

When compared to the full practice population, or to those that had consulted their general practice at all, the cohort of patients that had any recorded coding activity in 2019-2020 tended to skew towards female patients; towards white patients, and those least deprived. Age-wise, there was a higher relative preponderance of those aged 18-40. Though focussed on different types of general practice online service access (booking appointments, ordering prescriptions and viewing records, not online consultation system request or consultation), analysis of 2018 and 2019 GP patient survey data ^24,25^ previously showed analogous evidence of a strong deprivation gradient in awareness and use of services in favour of those least deprived as well as a reduction in awareness and use for those older than 75. Ethnicity was also associated with variability. A 2019 cross-sectional West Midlands self-administered survey also showed variation in use and awareness of those three services with demographics – namely lower levels with greater deprivation and with being male (awareness of prescriptions; awareness and use of online appointment booking) ^26^.

We also found early indications that patients with ‘eConsultation’ coding activity were more prevalent in those with a clinical history of asthma, depression or heart conditions than other patients with any overall GP consultation activity in the same period of time – in this case covering March 2020 to February 2021 inclusive. This increased activity may be reflecting expected increased general health utilisation in such cohorts but also adoption of potentially tailored e-consultation solutions and formularies for these long-term condition cohorts ^27^. However, it is important to note that the online consultation systems user profile may not be generalisable and may be very dependent on the practice-by-practice implementation model, type and design of online consultation system, communications and practice context, patient factors and change management support.

### Strengths and weaknesses

The key strength of the study relies on the scale and completeness of the underlying record-level EHR data. With OpenSAFELY-TPP, analysis can be run directly on the full dataset of raw, single-event clinical events, including tests, treatments, diagnostic events and other diagnostic and sociodemographic information, covering 40% of practices in England and equating to over 20 million patients, if considering those with a stable practice of registration over the study period.

When used alongside with OpenSAFELY-EMIS, this extends to about 99% of practices. Linkage to secondary care and mortality information is also inbuilt. In comparison with other general practice setting data sources, the CPRD dataset holds records on only a sample of patients across two databases, while the GPES dataset held by NHS Digital contains less data items for each individual patient. NHS England holds a number of record-level commissioning datasets, but these are primarily focussed on capturing secondary care activity, namely across inpatient, outpatient and emergency settings. NHS England open publications like GPAD can only give a high-level view of a proportion of activity, with no insight into clinical or sociodemographic factors, quality or the full workload of general practices. Further, they group coarsely all forms of e-consultation as mode of consultation (video, written) and do not provide an easy route to follow GP access types and pathways, though GPAD improvements are being rolled out – including on standardising appointment categories ^28^. Another key strength is the transparency and reproducibility of the analysis undertaken. All code for the platform, data management and analysis is shared openly on Github, allowing for peer review and re-usability under open licenses.

In terms of key limitations, as highlighted when describing the codelists in the Methods section, a data-driven approach is taken that relies on SNOMED codes and mapping to historical CTV3 hierarchy. Caution must be applied when interpreting the results as not all activity from use of the online consultation system is captured in the GP IT clinical system, the codes examined are not specific and it is unclear if they are being used to describe submissions made using an online consultation system, the consultation mode by written online message or another remote consultation modality, additionally practices vary in their coding practice and therefore use of the codes is unlikely to be consistent. The improvement in coding definition and quality will directly enable potential improvements in insight from OpenSAFELY (and from any other data assets leveraging EHR systems, like CPES and CPRD).

### Opportunities for future research

The first area to highlight relates to coding quality. By highlighting characteristics and gaps in coding approaches and activity, the insights from this study support work underway with system suppliers to improve coding guidelines and implementation. By regularly monitoring and reviewing the coding activity and reviewing it in context with further data sources and evidence on online consultation system implementation and use, coding quality could be proactively tracked and improved. This can be done through reduced burden for administrative and clinical staff, given that analysis can be executed in a single framework from re-executable code. Since OpenSAFELY encourages curation of thematic and open codelists, the existing codelist can be managed and updated in line with those developments and such that, when relevant, it becomes increasingly aligned with ongoing specification work for GPAD and the forthcoming NHS England online consultation system supplier collection being stood up.

Secondly, forward-looking analysis will be able to leverage insight brought by new upcoming SNOMED codes that better define and differentiate OCS submissions and online written consultations; that differentiate triage in terms of request type (clinical and admin), mode of consultation (written online consultation, video, telephone, face-to-face) and professional type, allowing for a more journey- or pathway-centric view of digital first primary care activity. The wider operationalisation of OpenSAFELY-EMIS also means that, alongside OpenSAFELY-TPP, studies will be able to draw more comprehensively on data encompassing over 99% of the English population ^29^.

The third area to highlight relates to future opportunities to use OpenSAFELY – alongside a refreshed curated codelist tailored to maximise the tracking of underlying patient activity, namely code ‘eConsultation’ - for large-scale cohort or longitudinal studies that help inform continuous improvement and evaluation on service utilisation and patient outcomes. Examples of research questions include the creation of an observatory of inequalities, where a strategic metric is chosen for tracking variation by a key protected characteristic; characterisation of the types of demand and requests submitted by online consultation system users; pathway analysis to support impact evaluation protocols (e.g. illustratively, impact of online access and comparisons of modes of consultations on continuity of care, rates of A&E attendance and admissions, secondary care referrals, general practice appointment utilisation by role and modality, re-utilisation and prescribing) ^6,7^. As for the assessment of real-world pilot studies in specific practices, including counterfactual analysis, by design outputs with disclosive practice identifiers cannot be extracted and querying on practice identifier is not yet straightforward given pseudonymisation. However, there is currently an ongoing cluster randomised controlled trial (RCT) project where such matching is being done in the background by TPP. Subject to ensuring such functionality would still comply with information governance and data protection - with appropriate processes, controls and safeguards in place – the ability to define a pilot (intervention) and counterfactual group in the OpenSAFELY study design could in future be extended to external collaborators through simplified platform functionality.

## Conclusions

Insights from this study increase understanding of the implementation and use of online consultation systems and written online consultations in terms of implementation, trend and variation. Alongside operational data and evaluation studies, this can support the evidence base around models of online consultation system implementation and differential patterns of access and uptake. Current gaps in coding practice are also highlighted and can therefore support conversations with practices, online consultation system suppliers and EHR suppliers on ensuring consistent and widespread coding practices. Further work that could be leveraged via OpenSAFELY includes key metric monitoring such as coding quality, coding activity or variation, as well as facilitation of large-scale impact evaluation studies to understand the types of demand and characteristics of online consultation system users and how the use of online consultation systems affects outcomes such as continuity of care, type and modality of general practice consultation use or unplanned urgent and emergency care.

## Data Availability

All data were linked, stored and analysed securely within the OpenSAFELY platform (https://opensafely.org/), a data analytics platform created with the approval of NHS England to address urgent COVID-19 research questions.
Patient data has been pseudonymised for analysis and linkage using industry standard cryptographic hashing techniques; all pseudonymised datasets transmitted for linkage onto OpenSAFELY are encrypted; access to the platform is via a virtual private network (VPN) connection, restricted to a small group of researchers; the researchers hold contracts with NHS England and only access the platform to initiate database queries and statistical models; all database activity is logged; only aggregate statistical outputs leave the platform environment following best practice for anonymisation of results such as statistical disclosure control for low cell counts. The aggregate statistical outputs are available online: https://github.com/opensafely/OS_OC_v001-research/tree/release-candidates/released_outputs. Similarly, the analytical codebase used in this study is openly available for inspection and re-use at https://github.com/opensafely/OS_OC_v001-research. All codelists used are openly available at OpenSAFELY Codelists (https://www.opencodelists.org/codelist/user/martinaf/online-consultations-snomed-v01/28bba9bc/).
Detailed pseudonymised patient data is potentially re-identifiable and therefore not shared. The queried pseudonymised records are available through the OpenSAFELY framework. Further details on our information governance can be found under information governance and ethics.

## Acknowledgements

We are very grateful for all the support received from the EMIS and TPP Technical Operations teams throughout this work, and for generous assistance from the information governance and database teams at NHS England (NHSE) Transformation Directorate. We would like to thank colleagues in the OpenSAFELY collaborative and in the NHS Transformation Directorate’s Digital Analytics and Research Team (DART) for the assistance and guidance in using the platform and in retrieving results. We would like to thank colleagues in the NHSE Primary Care Research Group for reviewing the manuscript.

## Author contributions

*Conceptualization: MF, OK, MB*.

*Data curation: MF, LF*.

*Formal analysis: MF*.

*Investigation: MF, BMK, OK, MB*.

*Funding acquisition:* The OpenSAFELY collaborative, BG.

*Platform process and approvals: AM*.

*Methodology: MF, BMK, OK, MB*.

*Resources: MF, BMK, AM, CEW, GH, LF, JP, PI, SB, SD, WH, BG*, The OpenSAFELY collaborative, *OK, MB*.

*Software:* The OpenSAFELY collaborative, *BMK, AM, GH, LF, JP, PI, SB, SD, WH, BG*.

*Supervision: OK, BMK*.

*Visualisation: MF*.

*Writing - original draft: MF, OK, MB*.

*Writing - review & editing: MF, BMK, AM, CEW, GH, LF, JP, PI, SB, SD, WH, BG, OK, MB*.

## Information Governance and Ethical Approval

NHS England is the data controller for OpenSAFELY-EMIS and OpenSAFELY-TPP; EMIS and TPP are the data processors; all study authors using OpenSAFELY have the approval of NHS England. This implementation of OpenSAFELY is hosted within EMIS and TPP environments which are accredited to the ISO 27001 information security standard and are NHS IG Toolkit compliant;^30,31^

Patient data has been pseudonymised for analysis and linkage using industry standard cryptographic hashing techniques; all pseudonymised datasets transmitted for linkage onto OpenSAFELY are encrypted; access to the platform is via a virtual private network (VPN) connection, restricted to a small group of researchers; the researchers hold contracts with NHS England and only access the platform to initiate database queries and statistical models; all database activity is logged; only aggregate statistical outputs leave the platform environment following best practice for anonymisation of results such as statistical disclosure control for low cell counts.^32^

The OpenSAFELY research platform adheres to the obligations of the UK General Data Protection Regulation (GDPR) and the Data Protection Act 2018. In March 2020, the Secretary of State for Health and Social Care used powers under the UK Health Service (Control of Patient Information) Regulations 2002 (COPI) to require organisations to process confidential patient information for the purposes of protecting public health, providing healthcare services to the public and monitoring and managing the COVID-19 outbreak and incidents of exposure; this sets aside the requirement for patient consent.^33^

Taken together, these provide the legal bases to link patient datasets on the OpenSAFELY platform. GP practices, from which the primary care data are obtained, are required to share relevant health information to support the public health response to the pandemic and have been informed of the OpenSAFELY analytics platform. This study was supported by Dr Minal Bakhai, National Clinical Director for Digital First Primary Care at NHS England, as senior sponsor.

### Guarantor

MF

## Supplementary material

Supplementary material for this paper is available.

## Footnotes

1 There are 5 online consultation system suppliers that are not currently submitting data to the national collection: Substrakt, Klinik, Askmynhs (Sensely), iPlato, At Medics. As of the 31^st^ of March 2021 there were 700 practices in England out of 6,621 that had online consultations systems but were not submitting data as part of the national collection.

2 Implementation supplier activity data on OC/VC collected under the Control of Patient Information notice under COVID. Figures retrieved 21/06/2021 from the *GP Online & Video Consultations Dashboard* on the FutureNHS Digital IPC Workspace [7]. The dashboard is available for NHS Organisations to use for the purposes of supporting implementation across Primary Care. Access requires authentication and authorisation.

3 A written online consultation is a two-way written exchange between a healthcare professional and a patient using an online medium (such as an online web platform or SMS).

5 cohortextractor.patients.with_gp_consultations() captures GP-patient interactions, whether in person or by phone/video call. The concept of a “consultation” in EHR systems is generally broader and might include things like updating a phone number with the receptionist. It captures events such as interactions and “consultations” differently from what is captured in the NHS Digital GP Appointment Data (GPAD) from GP Appointment Systems It might also - but will not necessarily - capture interactions from online consultation related events. In the analysis in this paper, the metric and shorthand terminology “GP consultation” or “GP-patient interaction” – whether in the methods, figures, tables or descriptions - will refer to that obtained via the OpenSAFELY EHR method above, with the relevant caveats.

6 Patient Online is an NHS England programme designed to support GP Practices to offer and promote online services to patients, including access to coded information in records, appointment booking and ordering of repeat prescriptions. Known as POMI (patient online management information), data are provided by GP system suppliers to NHS Digital on a monthly basis and published on the 15th working day each month pending no issues, otherwise as soon as possible thereafter.

