## Supplementary information (appendices) for "Primary care coding activity related to the use of online consultation systems or remote consulting: an analysis of 53 million peoples’ health records using OpenSAFELY"

#### Appendix 1. Further data - coding coverage and instance rates

The graph below provides information on TPP coverage (code ever used over two year period) broken down by region. Focussing on the eConsultation code, TPP practice coverage-wise, its coverage of use has been higher in the East and South East and lowest in the North East and Midlands. There may be considerations around practices with changed or multiple systems.

**Figure A1. Portion of TPP practices with *any* recorded activity for online consultation relevant codes *in general practice*** *(January 2019 - December 2020).* **Broken down by region.**

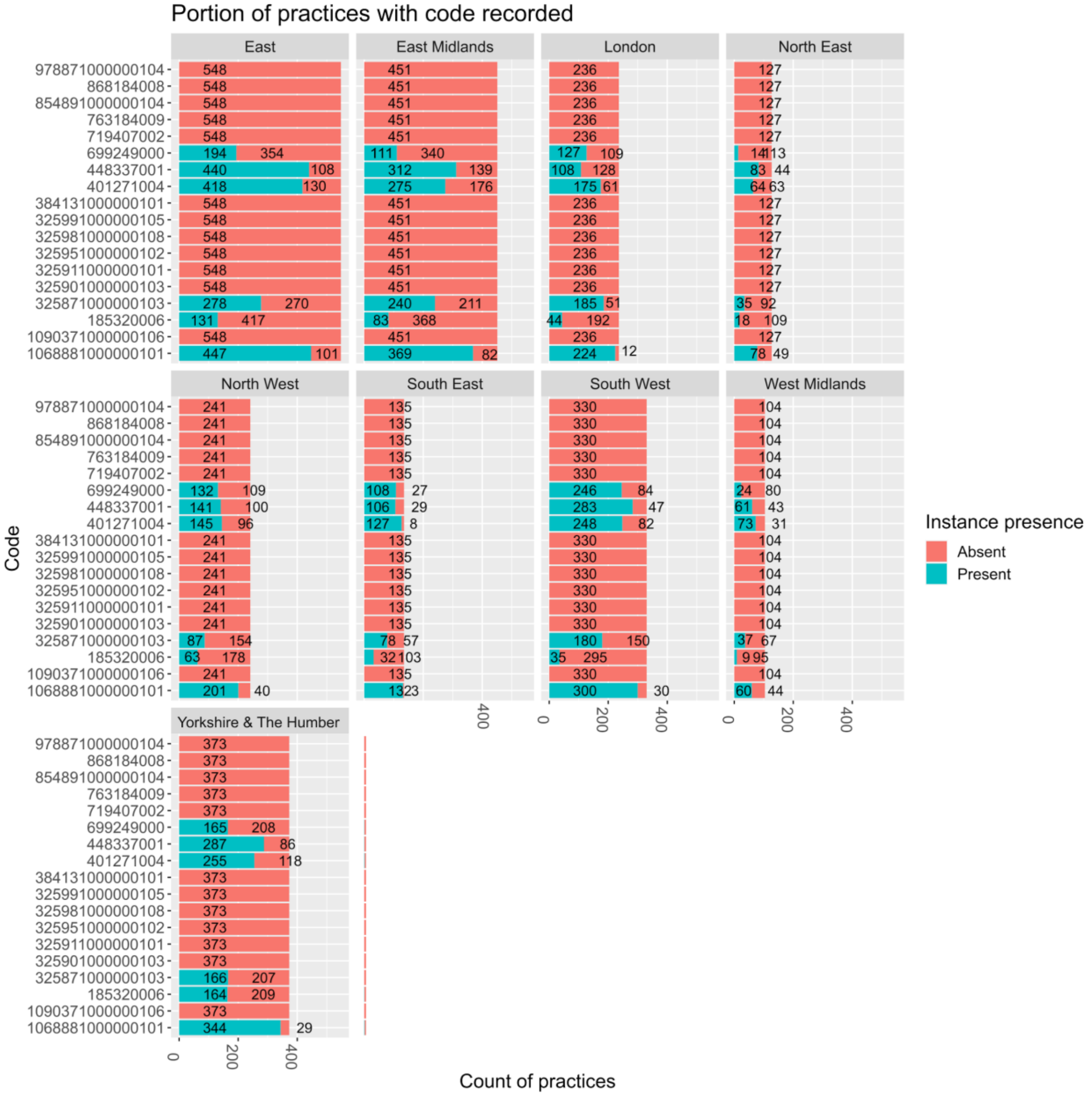

**Figure A2. Portion of TPP practices by region with *any* recorded activity for eConsultation code *in general practice*** *(January 2019 - December 2020). Numbers in white show absolute count of practices.*

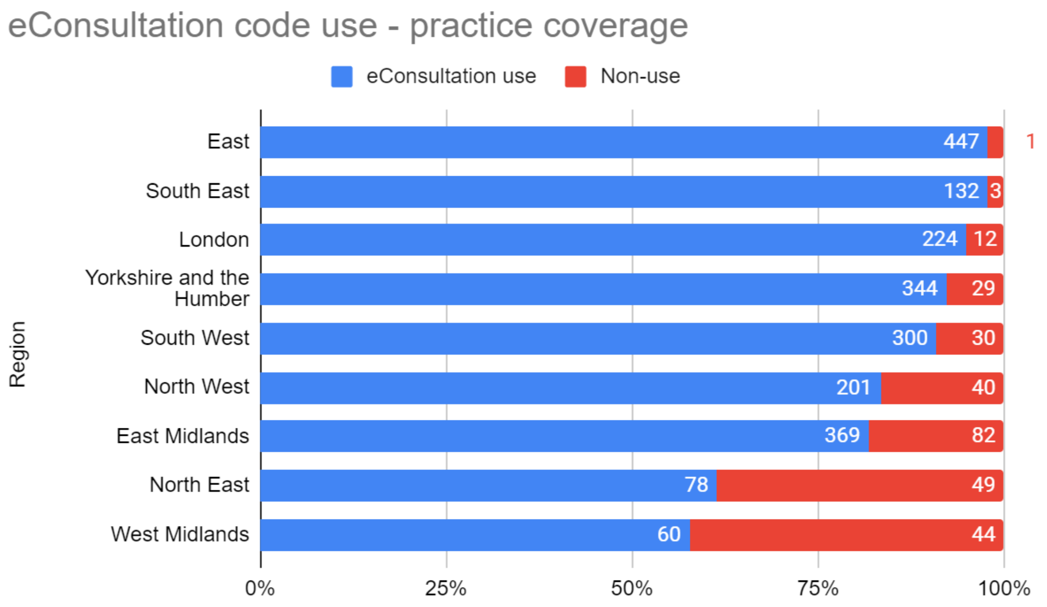

**Figure A3.** ***Monthly absolute counts of SNOMED codes in TPP general practice*** *(January 2019 - December 2020).* March 2020 indicated in pink. Note the different y axes (each plot scaled individually)

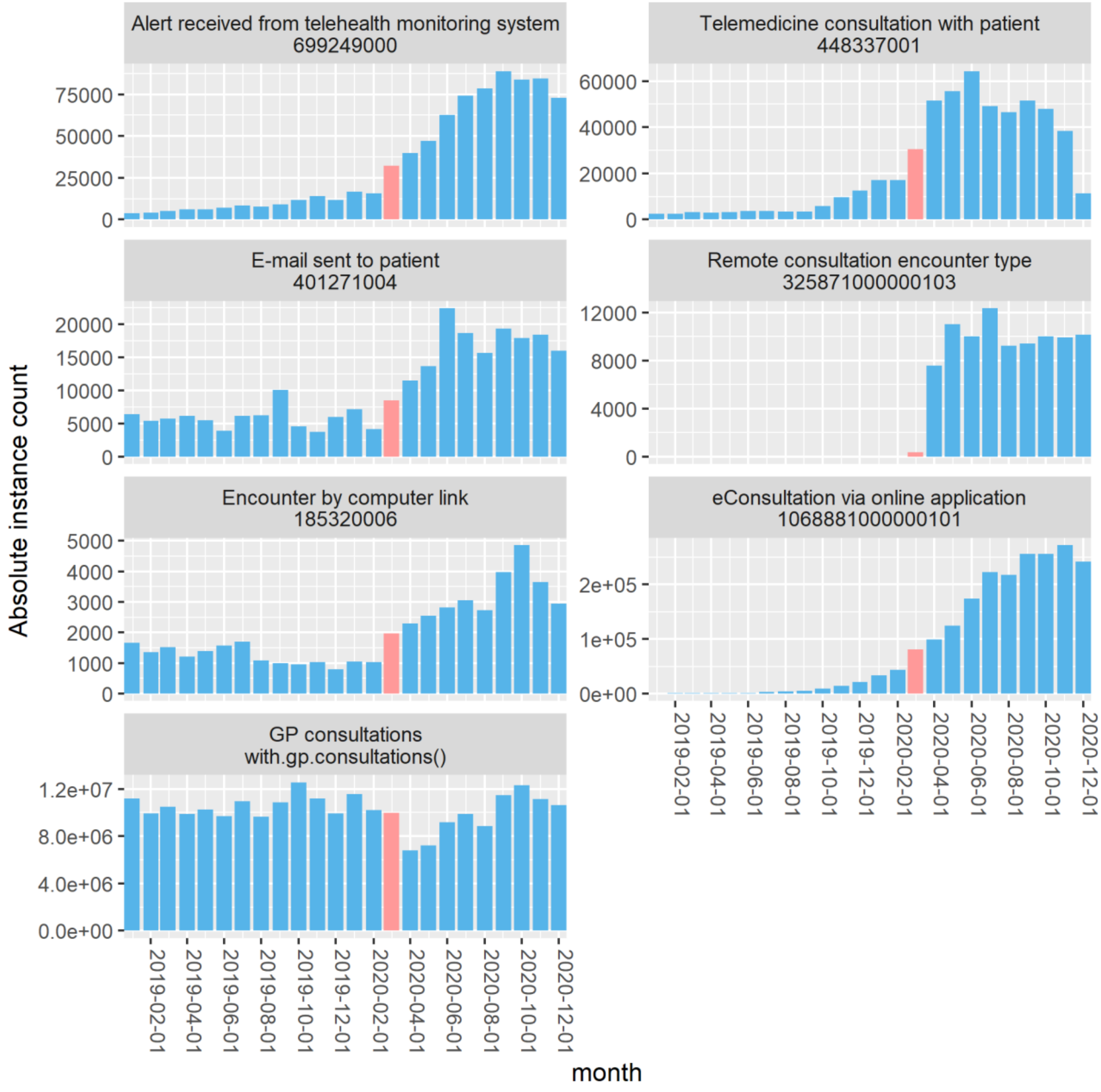

#### Appendix 2. Further data - TPP cohort sociodemographic characteristics

*Table A 1 Characteristics of the studied cohort, both overall and by a) patients without a GP consultation in 2019-2020; b) patients with a consultation.*

| **Characteristic** | **Overall,** N = 20,651,036[1] | **Had any GP consultation with.gp.consultations()** **NO**, N = 3,484,271[1] | **YES**, N = 17,166,765[1] | **p-value**[2] |
|---|---|---|---|---|
| **sex** | | | | <0.001 |
| Female | 10,260,731 (50%) | 1,087,898 (31%) | 9,172,833 (53%) | |
| Male | 10,389,976 (50%) | 2,396,331 (69%) | 7,993,645 (47%) | |
| Other/Unknown | 329 (<0.1%) | 42 (<0.1%) | 287 (<0.1%) | |
| **age** | 41 (22, 59) | 30 (15, 43) | 44 (24, 61) | <0.001 |
| **age group** | | | | <0.001 |
| (0,18] | 4,298,691 (21%) | 1,019,206 (29%) | 3,279,485 (19%) | |
| (18,40] | 5,738,142 (28%) | 1,442,431 (41%) | 4,295,711 (25%) | |
| (40,50] | 2,842,130 (14%) | 488,196 (14%) | 2,353,934 (14%) | |
| (50,60] | 2,913,528 (14%) | 336,154 (9.7%) | 2,577,374 (15%) | |
| (60,70] | 2,269,212 (11%) | 131,588 (3.8%) | 2,137,624 (13%) | |
| (70,80] | 1,673,588 (8.2%) | 46,058 (1.3%) | 1,627,530 (9.6%) | |
| (80,Inf] | 746,742 (3.6%) | 17,458 (0.5%) | 729,284 (4.3%) | |
| Unknown | 169,003 | 3,180 | 165,823 | |
| **ethnicity** | | | | <0.001 |
| Asian | 1,252,414 (6.1%) | 256,846 (7.4%) | 995,568 (5.8%) | |
| Black | 412,399 (2.0%) | 93,541 (2.7%) | 318,858 (1.9%) | |
| Mixed | 249,470 (1.2%) | 60,171 (1.7%) | 189,299 (1.1%) | |
| Other | 6,026,577 (29%) | 1,251,395 (36%) | 4,775,182 (28%) | |
| White | 12,710,176 (62%) | 1,822,318 (52%) | 10,887,858 (63%) | |
| **living alone** | 5,783,003 (28%) | 1,049,450 (30%) | 4,733,553 (28%) | <0.001 |
| **region** | | | | <0.001 |
| East | 4,823,404 (23%) | 809,148 (23%) | 4,014,256 (23%) | |
| East Midlands | 3,618,902 (18%) | 581,969 (17%) | 3,036,933 (18%) | |
| London | 1,340,024 (6.5%) | 399,214 (11%) | 940,810 (5.5%) | |
| North East | 963,807 (4.7%) | 151,347 (4.3%) | 812,460 (4.7%) | |
| North West | 1,843,088 (8.9%) | 255,636 (7.3%) | 1,587,452 (9.2%) | |
| South East | 1,357,871 (6.6%) | 238,663 (6.9%) | 1,119,208 (6.5%) | |
| South West | 2,838,383 (14%) | 427,567 (12%) | 2,410,816 (14%) | |
| West Midlands | 861,670 (4.2%) | 161,955 (4.6%) | 699,715 (4.1%) | |
| Yorkshire & The Humber | 2,997,813 (15%) | 457,756 (13%) | 2,540,057 (15%) | |
| Unknown | 6,074 | 1,016 | 5,058 | |

| | | | | |
|---|---|---|---|---|
| **deprivation quintile** | | | | <0.001 |
| Q1 (least) | 4,157,772 (20%) | 781,369 (23%) | 3,376,403 (20%) | |
| Q2 | 4,032,329 (20%) | 735,329 (21%) | 3,297,000 (20%) | |
| Q3 | 4,259,619 (21%) | 714,307 (21%) | 3,545,312 (21%) | |
| Q4 | 4,052,737 (20%) | 642,990 (19%) | 3,409,747 (20%) | |
| Q5 (most) | 3,796,821 (19%) | 558,209 (16%) | 3,238,612 (19%) | |
| Unknown | 351,758 | 52,067 | 299,691 | |
| **rural urban** | | | | <0.001 |
| Other | 328,860 (1.6%) | 47,257 (1.4%) | 281,603 (1.6%) | |
| Rural | 4,113,110 (20%) | 535,061 (15%) | 3,578,049 (21%) | |
| Urban | 16,209,066 (78%) | 2,901,953 (83%) | 13,307,113 (78%) | |
| **care home** | | | | <0.001 |
| Yes | 37,137 (0.2%) | 2,057 (<0.1%) | 35,080 (0.2%) | |
| Non | 20,613,899 (100%) | 3,482,214 (100%) | 17,131,685 (100%) | |

[1] n (%); Median (IQR)

[2] Pearson's Chi-squared test; Wilcoxon rank sum test

#### Ethnicity

OC coding activity is lower for non-white patients, both in terms of coverage and instance rates. Asian and Black patients register the lowest rates and coverage. Though these differential patterns are also present in GP consultation rates and coverage, they are not as pronounced.

It should be noted that about 6M patients had no ethnicity recorded, or an explicit 'Other' ethnicity, based on GP clinical coding.

*Table A 2 Ethnicity breakdown. Online consultation coding instance rates (per 1,000 registered practice patient population) and population coverage, considering the period of 2019-2020 and patients registered with a single practice. Figures for GP consultations are also given for context.*

| ethnicity | Cohort * | GP consultation rate | | GP consultation coverage | | OC instance rate | | OC instance coverage | |
|---|---|---|---|---|---|---|---|---|---|
| Asian | 1,252,400 | 11.15 | 0.91 | 79.5% | 0.93 | 0.09 | 0.53 | 3.4% | 0.60 |
| Black | 412,400 | 10.06 | 0.82 | 77.3% | 0.90 | 0.09 | 0.53 | 3.4% | 0.60 |
| Mixed | 249,500 | 8.92 | 0.73 | 75.9% | 0.89 | 0.12 | 0.71 | 4.3% | 0.75 |
| Other | 6,026,600 | 8.72 | 0.71 | 79.2% | 0.92 | 0.14 | 0.82 | 4.8% | 0.84 |
| White | 12,710,200 | 12.2 | 1.00 | 85.7% | 1.00 | 0.17 | 1.00 | 5.7% | 1.00 |

* rounded to nearest 100

Gray figures are ratio vs White

#### Deprivation

OC coding activity is lower for the most deprived patients, both in terms of coverage and 2019-2020 instance rates. This deprivation pattern (direction-wise) is also seen in terms of GP consultation coverage, but not for GP consultation rates. Overall GP consultation rates are higher among the most deprived.

*Table A 3 Deprivation breakdown. Online consultation coding instance rates (per 1,000 registered practice patient cohort population) and population coverage, considering the period of 2019-2020 and patients registered with a single practice. Figures for GP consultations are also given for context.*

| deprivation | Cohort * | GP consultation rate | | GP consultation coverage | | OC instance rate | | OC instance coverage | |
|---|---|---|---|---|---|---|---|---|---|
| 1 (most) | 4,157,800 | 11.32 | 1.06 | 81.2% | 0.95 | 0.12 | 0.75 | 4.0% | 0.70 |
| 2 | 4,032,300 | 11.05 | 1.04 | 81.8% | 0.96 | 0.15 | 0.94 | 5.2% | 0.90 |
| 3 | 4,259,600 | 11.13 | 1.05 | 83.2% | 0.98 | 0.15 | 0.94 | 5.5% | 0.96 |
| 4 | 4,052,700 | 10.94 | 1.03 | 84.1% | 0.99 | 0.17 | 1.06 | 5.8% | 1.01 |
| 5 (least) | 3,796,800 | 10.64 | 1.00 | 85.3% | 1.00 | 0.16 | 1.00 | 5.8% | 1.00 |

* rounded to nearest 100

Gray figures are ratio vs least deprived

#### Age-Sex

OC coding activity and coverage has been higher for female patients. Coverage and coding activity has been highest for females aged 18-40 (over the two years, there were 0.29 coding events per 1,000 cohort registered population and 8.6% of the cohort had at least one instance), followed by females aged 40-50, 50-60 and 60-70, in this order.

*Table A 4 Age and sex breakdown. Online consultation coding instance rates (per 1,000 practice patient cohort population) and population coverage, considering the period of 2019-2020 and patients registered with a single practice. Figures for GP consultations are also given for context.*

| Age group | Sex | Cohort * | GP consultation rate | GP consultation coverage | OC instance rate | OC instance coverage |
|---|---|---|---|---|---|---|
| (0,18] | Female | 2,079,900 | 5.64 | 78.9% | 0.10 | 3.9% |
| (0,18] | Male | 2,218,700 | 4.21 | 73.8% | 0.07 | 3.0% |
| (18,40] | Female | 2,774,900 | 13.82 | 87.5% | 0.29 | 8.6% |
| (18,40] | Male | 2,963,100 | 5.08 | 63.0% | 0.10 | 3.8% |
| (40,50] | Female | 1,388,200 | 13.50 | 91.8% | 0.26 | 8.1% |
| (40,50] | Male | 1,453,900 | 7.55 | 74.3% | 0.12 | 4.4% |
| (50,60] | Female | 1,440,400 | 14.09 | 92.9% | 0.22 | 7.3% |
| (50,60] | Male | 1,473,200 | 10.20 | 84.1% | 0.13 | 5.0% |
| (60,70] | Female | 1,157,800 | 16.23 | 95.4% | 0.16 | 5.7% |
| (60,70] | Male | 1,111,400 | 14.71 | 92.9% | 0.14 | 5.3% |
| (70,80] | Female | 889,400 | 20.47 | 97.5% | 0.12 | 4.3% |
| (70,80] | Male | 784,200 | 19.61 | 97.0% | 0.12 | 4.7% |
| 80+ | Female | 447,700 | 23.71 | 97.9% | 0.11 | 4.0% |
| 80+ | Male | 299,100 | 24.49 | 97.4% | 0.11 | 4.1% |

* rounded to nearest 100

#### Learning and intellectual disabilities

OC coding activity coverage is similar for those with and without disability, at about 5% (though slightly higher for those without disability). This contrasts with GP consultation coverage, which was higher for those with disability than those without (87.7% vs 83.0%). Patterns may not be statistically significant.

For this work, the presence of a disability was defined by identifying patients with codes from codelists related to QOF register learning disabilities and intellectual disabilities. Physical disabilities were not covered.

*Table A 5 Disability flag breakdown. Online consultation coding instance rates (per 1,000 practice patient population) and population coverage, considering the period of 2019-2020 and patients registered with a single practice. Figures for GP consultations are also given for context.*

| Disability | Cohort * | GP consultation rate | GP consultation coverage | OC instance rate | OC instance coverage |
|---|---|---|---|---|---|
| No | 20,281,600 | 11.0 | 83.0% | 0.15 | 5.3% |
| Yes | 369,500 | 12.4 | 87.7% | 0.15 | 5.1% |

* rounded to nearest 100

#### Region and Rurality

A breakdown by Region and rurality is given below (areas classed as ‘Other’ rurality-wise were excluded).

*Table A 6 Region and rurality breakdown. Online consultation coding instance rates (per 1,000 practice patient population) and population coverage, considering the period of 2019-2020 and patients registered with a single practice. Figures for GP consultations are also given for context.*

| Region | Rurality # | Cohort * | GP consultation rate | GP consultation coverage | OC instance rate | OC instance coverage |
|---|---|---|---|---|---|---|
| East | Rural | 1,202,300 | 11.76 | 86.3% | 0.10 | 4.1% |
| | Urban | 3,560,900 | 10.56 | 82.2% | 0.10 | 4.2% |
| East Midlands | Rural | 799,700 | 12.26 | 87.9% | 0.24 | 5.1% |
| | Urban | 2,779,200 | 11.21 | 82.8% | 0.17 | 4.2% |
| London | Rural | 1,400 | 10.05 | 64.4% | 0.09 | 5.0% |
| | Urban | 1,323,800 | 9.15 | 70.2% | 0.11 | 4.7% |
| North East | Rural | 97,400 | 12.59 | 87.5% | <0.01 | 0.4% |
| | Urban | 858,300 | 11.35 | 83.9% | <0.01 | 0.4% |
| North West | Rural | 417,600 | 12.71 | 88.7% | 0.16 | 6.1% |
| | Urban | 1,395,000 | 10.79 | 85.3% | 0.19 | 6.7% |
| South East | Rural | 265,000 | 10.94 | 85.8% | 0.18 | 7.8% |
| | Urban | 1,070,200 | 10.12 | 81.6% | 0.26 | 9.2% |
| South West | Rural | 887,400 | 11.94 | 86.5% | 0.17 | 6.9% |
| | Urban | 1,913,200 | 10.98 | 84.2% | 0.31 | 9.7% |
| West Midlands | Rural | 21,100 | 9.47 | 81.5% | <0.01 | 0.2% |
| | Urban | 834,800 | 9.43 | 81.2% | 0.06 | 2.5% |

* rounded to nearest 100
### excludes 'Other'

#### Care home status

As with GP consultation rates and coverage, OC coding activity coverage and instance rates are higher for care home residents than the remaining population. About 8,700 OC coding instances have been identified for care home residents. Subsequent analysis could potentially explore these patterns focussing only on the elderly population.

Care home status was assessed using TPP's functionality and the full detail on this methodology, its strengths and limitations can be read in the OpenSAFELY short data report published on Wellcome Open Research [16].

*Table A 7 Breakdown for care home residency. Online consultation coding instance rates (per 1,000 practice patient population) and population coverage, considering the period of 2019-2020 and patients registered with a single practice. Figures for GP consultations are also given for context.*

| Care home | Cohort * | GP consultations * | GP consultation rate | GP consultation coverage | OC instances * | OC instance rate | OC instance coverage |
|---|---|---|---|---|---|---|---|
| Yes | 37,100 | 827,500 | 22.28 | 94.5% | 8,700 | 0.23 | 0.07 |
| Non | 20,613,900 | 227,134,200 | 11.02 | 83.1% | 3,137,100 | 0.15 | 0.05 |

* rounded to nearest 100

### Appendix 3. TPP cohort clinical history (eConsultation)

*Table A 8 Clinical history characteristics of the cohort with eConsultation code recorded in March 2020-February 2021. Comparison against remaining population in those practices.*

| **Clinical history (pre-March 2019)** | **Overall**, N = 14,677,783[1] | **Had eConsultation code instance in Mar20-Feb21** | | |
|---|---|---|---|---|
| | | No, N = 13,860,236[1] | Yes, N = 817,547[1] | p-value[2] |
| history_hypertension | 2,349,943 (16%) | 2,219,265 (16%) | 130,678 (16%) | 0.5 |
| history_asthma | 2,153,773 (15%) | 1,988,905 (14%) | 164,868 **(20%)** | <0.001 |
| history_osteoarthritis | 1,634,638 (11%) | 1,541,658 (11%) | 92,980 (11%) | <0.001 |
| history_depression | 2,266,604 (15%) | 2,080,081 (15%) | 186,523 **(23%)** | <0.001 |
| history_diabetes | 1,054,747 (7.2%) | 996,681 (7.2%) | 58,066 (7.1%) | 0.003 |
| history_chronic_heart_disease | 661,437 (4.5%) | 628,058 (4.5%) | 33,379 (4.1%) | <0.001 |
| history_cancer | 577,276 (3.9%) | 543,264 (3.9%) | 34,012 **(4.2%)** | <0.001 |
| history_atrial_fibrillation | 294,501 (2.0%) | 279,583 (2.0%) | 14,918 (1.8%) | <0.001 |
| history_stroke | 209,562 (1.4%) | 199,702 (1.4%) | 9,860 (1.2%) | <0.001 |
| history_chronic_respiratory_disease | 423,574 (2.9%) | 400,087 (2.9%) | 23,487 (2.9%) | 0.5 |
| history_peripheral_arterial_disease | 93,554 (0.6%) | 89,524 (0.6%) | 4,030 (0.5%) | <0.001 |
| history_heart_failure | 157,763 (1.1%) | 149,755 (1.1%) | 8,008 (1.0%) | <0.001 |
| history_chronic_kidney_disease | 14,087 (<0.1%) | 13,120 (<0.1%) | 967 (0.1%) | <0.001 |
| history_serious_mental_illness | 127,592 (0.9%) | 120,010 (0.9%) | 7,582 (0.9%) | <0.001 |
| econsult_pre_had [3] | 60,082 (0.4%) | 21,896 (0.2%) | 38,186 (4.7%) | |
| gp_consult_pre_had [3] | 10,592,688 (72%) | 9,879,901 (71%) | 712,787 (87%) | |
| gp_consult_post_had [3] | 9,811,243 (67%) | 9,018,200 (65%) | 793,043 (97%) | |

[1] n (%)

[2] Pearson's Chi-squared test

[3] 'Pre' refers to Mar19-Feb20. 'Post' refers to Mar20-Feb21

*Table A 9 Clinical history characteristics of the cohort with eConsultation code recorded in March 2019-February 2020. Comparison against remaining population in those practices that had GP consultation recorded.*

| **Clinical history (pre-March 2019)#** | **Overall**, N = 3,329,385[1] | **Had eConsultation code instance in Mar19-Feb20 (among those with an eConsultation/GP consultation)** | | |
|---|---|---|---|---|
| | | **No**, N = 3,269,239[1] | **Yes**, N = 60,146[1] | p-value[2] |
| history_hypertension | 692,860 (21%) | 683,048 (21%) | 9,812 (16%) | <0.001 |
| history_asthma | 561,217 (17%) | 548,600 (17%) | 12,617 (21%) | <0.001 |
| history_osteoarthritis | 482,702 (14%) | 475,669 (15%) | 7,033 (12%) | <0.001 |
| history_depression | 642,643 (19%) | 626,765 (19%) | 15,878 (26%) | <0.001 |
| history_diabetes | 300,541 (9.0%) | 296,013 (9.1%) | 4,528 (7.5%) | <0.001 |
| history_chronic_heart_disease | 195,284 (5.9%) | 192,640 (5.9%) | 2,644 (4.4%) | <0.001 |
| history_cancer | 168,067 (5.0%) | 165,543 (5.1%) | 2,524 (4.2%) | <0.001 |
| history_atrial_fibrillation | 89,142 (2.7%) | 88,042 (2.7%) | 1,100 (1.8%) | <0.001 |
| history_stroke | 61,494 (1.8%) | 60,818 (1.9%) | 676 (1.1%) | <0.001 |
| history_chronic_respiratory_disease | 130,723 (3.9%) | 128,830 (3.9%) | 1,893 (3.1%) | <0.001 |
| history_peripheral_arterial_disease | 29,255 (0.9%) | 28,924 (0.9%) | 331 (0.6%) | <0.001 |
| history_heart_failure | 46,687 (1.4%) | 46,117 (1.4%) | 570 (0.9%) | <0.001 |
| history_chronic_kidney_disease | 4,034 (0.1%) | 3,964 (0.1%) | 70 (0.1%) | 0.7 |
| history_serious_mental_illness | 35,964 (1.1%) | 35,367 (1.1%) | 597 (1.0%) | 0.036 |

| | | | |
|---|---|---|---|
| econsult_post_had [3] | 358,272 (11%) | 320,086 (9.8%) | 38,186 (63%) |
| gp_consult_pre_had [3] | 3,328,479 (100%) | 3,269,239 (100%) | 59,240 (98%) |
| gp_consult_post_had [3] | 2,676,230 (80%) | 2,623,722 (80%) | 52,508 (87%) |

[1] n (%)

[2] Pearson's Chi-squared test

[3] ‘Pre’ refers to Mar19-Feb20. ‘Post’ refers to Mar20-Feb21

#### Appendix 4 - Coding activity prevalence [EMIS]

Given the recent extension of OpenSAFELY to EMIS as well, a more fixed-scope overview of OC-related code use in EMIS in 2019-2020 was conducted, including: the individual codes in use, the coding activity volumes and population-adjusted rates.

All six codes in use in TPP were also in use in EMIS (Table A 11). Additionally, the following three codes were also found in at least five practices: Remote assessment encounter type (325951000000102) in five practices, Consultation via multimedia encounter type (325911000000101) in 271 (7%) of practices and Telehealth encounter type (854891000000104) in eight practices.

The three most common codes in terms of instances over two years per 1,000 EMIS population were in order: E-mail sent to patient (401271004) - 174.0 per 1,000; eConsultation via online application (1068881000000101) - 41.1 per 1,000; Alert received from telehealth monitoring system (699249000) - 37.5 per 1,000.

Table A 11 provides a comparison of the findings for TPP and EMIS in terms of instance rates and the number of practices (based on patient January 2019 registration) where patients activity for that code in 2019-2020 (note that for TPP this differs from Figure 2 'practice coverage', where patients are tracked month-by-month in terms of practice of registration).

*Table A 10 EMIS cohort over 2019-2020 (two-year period): code-by-code proportion practices with code, number of instances over two-year period, and population rates given both by 1,000 EMIS cohort population and covered practice population respectively. Practices as of 1 January 2019 (n=3,872). 30,542,038 patients were in scope.*

| Code | % of Jan19 practices with code | % Practice s | Number of instances (2019-2020) | Instance rate (per 1,000 EMIS pop.) | Instance rate (per 1,000 covered practice pop.) |
|---|---|---|---|---|---|
| eConsultation via online application (1068881000000100) | 1977 | 51.1% | 1,255,076 | 41.1 | 74.8 |
| Referral to remote triage and advice service (1090371000000100) | * | * | * | * | * |
| Encounter by computer link (185320006) | 298 | 7.7% | 118,022 | 3.9 | 36.7 |
| Remote consultation encounter type (325871000000103) | 278 | 7.2% | 10,565 | 0.3 | 3.6 |
| Remote non-verbal consultation encounter type (325901000000103) | * | * | * | * | * |
| Consultation via multimedia encounter type (325911000000101) | 271 | 7.0% | 119,163 | 3.9 | 43.9 |
| Remote assessment encounter type (325951000000102) | 5 | 0.1% | 20 | 0.0 | 0.3 |
| Remote non-verbal assessment encounter type (325981000000108) | * | * | * | * | * |
| Assessment via multimedia encounter type (325991000000105) | * | * | * | * | * |
| Remote encounter type (384131000000101) | * | * | * | * | * |
| E-mail sent to patient (401271004) | 3062 | 79.1% | 5,314,194 | 174.0 | 205.4 |
| Telemedicine consultation with patient (448337001) | 15 | 0.4% | 82 | 0.0 | 0.4 |
| Alert received from telehealth monitoring system (699249000) | 504 | 13.0% | 1,145,020 | 37.5 | 239.8 |
| Remote non-verbal consultation (719407002) | * | * | * | * | * |
| Telepractice consultation (763184009) | * | * | * | * | * |
| Telehealth encounter type (854891000000104) | 8 | 0.2% | 12 | 0.0 | 0.1 |
| Telemedicine consultation with provider (868184008) | * | * | * | * | * |
| Consultation via multimedia (978871000000104) | * | * | * | * | * |
| Any of the codes (OCall) | 3427 | 88.5% | 7,962,694 | 260.7 | 282.2 |

** redacted as less than five or no practices had the code*

*Table A 11 EMIS and TPP cohort over 2019-2020 (two-year period): proportion of practices with each SNOMED code, number of instances over two-year period per respective 1,000 EHR cohort population. Practices as of 1 January 2019, with patient activity within the 2019-2020 period assigned to their practice of registration on 1 January 2019 (for TPP this will therefore differ from Figure 2 'coverage', where patients are tracked month-by-month registration-wise). 23,433,739 patients were in scope for TPP.*

| Code | % of Jan19 practices with code | | Code instance rate (per 1,000 EHR population) | |
|---|---|---|---|---|
| | EMIS | TPP | EMIS | TPP |
| eConsultation via online application (1068881000000100) | 51.1% | 98.0% | 41.1 | 82.9 |
| Referral to remote triage and advice service (1090371000000100) | * | * | * | * |
| Encounter by computer link (185320006) | 7.7% | 38.0% | 3.9 | 4.2 |
| Remote consultation encounter type (325871000000103) | 7.2% | 63.3% | 0.3 | 28.8 |
| Remote non-verbal consultation encounter type (325901000000103) | * | * | * | * |
| Consultation via multimedia encounter type (325911000000101) | 7.0% | * | 3.9 | * |
| Remote assessment encounter type (325951000000102) | 0.1% | * | 0.0 | * |
| Remote non-verbal assessment encounter type (325981000000108) | * | * | * | * |
| Assessment via multimedia encounter type (325991000000105) | * | * | * | * |
| Remote encounter type (384131000000101) | * | * | * | * |
| E-mail sent to patient (401271004) | 79.1% | 89.4% | 174.0 | 11.7 |
| Telemedicine consultation with patient (448337001) | 0.4% | 88.5% | 0.0 | 23.2 |
| Alert received from telehealth monitoring system (699249000) | 13.0% | 85.2% | 37.5 | 34.2 |
| Remote non-verbal consultation (719407002) | * | * | * | * |
| Telepractice consultation (763184009) | * | * | * | * |
| Telehealth encounter type (854891000000104) | 0.2% | * | 0.0 | * |
| Telemedicine consultation with provider (868184008) | * | * | * | * |
| Consultation via multimedia (978871000000104) | * | * | * | * |
| Any of the codes (OCall) | 88.5% | 100% | 260.7 | 152.1 |

** redacted as less than five or no practices had the code*
